# Pan-cancer analysis of pre-diagnostic blood metabolite concentrations in the European Prospective Investigation into Cancer and Nutrition

**DOI:** 10.1101/2022.04.11.22273693

**Authors:** Marie Breeur, Pietro Ferrari, Laure Dossus, Mazda Jenab, Mattias Johansson, Sabina Rinaldi, Ruth C. Travis, Mathilde His, Tim J. Key, Julie A. Schmidt, Kim Overvad, Anne Tjønneland, Cecilie Kyrø, Joseph A. Rothwell, Nasser Laouali, Gianluca Severi, Rudolf Kaaks, Verena Katzke, Matthias B. Schulze, Fabian Eichelmann, Domenico Palli, Sara Grioni, Salvatore Panico, Rosario Tumino, Carlotta Sacerdote, Bas Bueno-de-Mesquita, Karina Standahl Olsen, Torkjel Manning Sandanger, Therese Haugdahl Nøst, J. Ramón Quirós, Catalina Bonet, Miguel Rodríguez Barranco, María-Dolores Chirlaque, Eva Ardanaz, Malte Sandsveden, Jonas Manjer, Linda Vidman, Matilda Rentoft, David Muller, Kostas Tsilidis, Alicia K. Heath, Hector Keun, Jerzy Adamski, Pekka Keski-Rahkonen, Augustin Scalbert, Marc J. Gunter, Vivian Viallon

## Abstract

**Background:** Epidemiological studies of associations between metabolites and cancer risk have typically focused on specific cancer types separately. Here, we designed a multivariate pan-cancer analysis to identify metabolites potentially associated with multiple cancer types, while also allowing the investigation of cancer type-specific associations.

**Methods:** We analyzed targeted metabolomics data available for 5,828 matched case-control pairs from cancer-specific case-control studies on breast, colorectal, endometrial, gallbladder, kidney, localized and advanced prostate cancer, and hepatocellular carcinoma nested within the European Prospective Investigation into Cancer and Nutrition (EPIC) cohort. From pre-diagnostic blood levels of an initial set of 117 metabolites, 33 cluster representatives of strongly correlated metabolites, and 17 single metabolites were derived by hierarchical clustering. The mutually adjusted associations of the resulting 50 metabolites with cancer risk were examined in penalized conditional logistic regression models adjusted for body mass index, using the data shared lasso penalty.

**Results:** Out of the 50 studied metabolites, *(i)* six were inversely associated with risk of most cancer types: glutamine, butyrylcarnitine, lysophosphatidylcholine a C18:2 and three clusters of phosphatidylcholines (PCs); *(ii)* three were positively associated with most cancer types: proline, decanoylcarnitine and one cluster of PCs; and *(iii)* 10 were specifically associated with particular cancer types, including histidine that was inversely associated with colorectal cancer risk, and one cluster of sphingomyelins that was inversely associated with risk of hepatocellular carcinoma and positively with endometrial cancer risk.

**Conclusions:** These results could provide novel insights for the identification of pathways for cancer development, in particular those shared across different cancer types.

## Background

Metabolomics allows the simultaneous measurement of a large variety of compounds present in biological samples, such as human blood^1,2^. Circulating metabolite levels can reflect both endogenous and exogenous processes, providing a snapshot of biological activity^3,4^. As a result, metabolomics may facilitate the identification of biological mechanisms involved in the development of chronic diseases. For example, prior metabolomics studies have identified metabolites associated with the risk of various chronic conditions, including type-2 diabetes (T2D)^5–7^, cardiovascular diseases (CVD)^8–10^, and different site-specific cancers, including cancers of the breast^11^, prostate^12,13^, endometrium^14^, kidney^15^, colorectum^16–18^, hepatocellular carcinoma (HCC)^19^, and others^20,21^.

Several shared biological mechanisms are known to underlie multiple chronic diseases. Obesity, physical inactivity and adherence to a Western-type diet, as well as chronic inflammation and insulin resistance, are recognized risk factors for cardio-metabolic diseases, including T2D, CVD, and several site-specific cancers^22–24^. Metabolomics may help uncover novel etiological mechanisms that are common to several chronic diseases as well as those that are disease-specific. One recent study identified metabolites associated with the risk of multimorbidity, defined as the simultaneous presence of multiple chronic conditions within one individual. Focusing on a pre-defined panel of metabolites, a targeted metabolomics study of breast, prostate and colorectal cancers in a German population found that circulating levels of the phosphatidylcholine PC ae C30:0 and several lysophosphatidylcholines, including lysoPC a C18:0, were predictive of the development of any of these three cancers^25^, suggesting that some etiological mechanisms could be shared across multiple cancer types.

In this work, we extended this concept by leveraging targeted metabolomics data available within nested case-control studies on eight cancer types (breast, colorectal, endometrial, gallbladder and biliary tract, kidney, localized prostate and advanced prostate cancers, and HCC) previously acquired in the European Prospective Investigation into Cancer and Nutrition (EPIC)^11,12,14,15,19^. The data shared lasso^26–28^, a penalized multivariate approach specifically designed for the investigation of a set of shared risk factors across different disease outcomes, was used to carry out a multivariate pan-cancer analysis to identify mutually adjusted metabolites associated with cancer risk and to identify those metabolites with consistent or heterogeneous patterns of associations across the eight cancer types.

## Methods

### Study population

EPIC is an ongoing multicentric prospective study with over 500,000 men and women recruited between 1992 and 2000 from 23 centers in 10 European countries^29^, originally designed to study the relationship between diet and cancer risk. Incident cancer cases were identified through a combination of methods, including health insurance records, cancer and pathology registries and active follow-up through study participants and their next-of-kin. At recruitment, information on diet and lifestyle was collected via self-administered questionnaires. Blood samples were collected from around 386,000 participants according to a standardized protocol. In France, Germany, Greece, Italy, the Netherlands, Norway, Spain, and the UK, serum (except in Norway), plasma, erythrocytes, and buffy coat aliquots were stored in liquid nitrogen (− 196 °C) in a centralized biobank at the International Agency for Research on Cancer (IARC). In Denmark, blood fractions were stored locally in the vapor phase of liquid nitrogen containers (− 150 °C), and in Sweden, they were stored locally at − 80 °C in standard freezers. Fasting was not required.

Our analyses used a set of metabolomics measurements from 15,948 EPIC participants from seven cancer-specific matched case-control studies nested within EPIC (Table 1). In each study, each case was matched to one control selected among cancer-free participants (other than non-melanoma skin cancer) by risk set sampling, using matching factors that included study center, sex, age at blood collection, time of the day of blood collection, fasting status, and use of exogenous hormones for women. All participants provided written informed consent to participate in the EPIC study. The cancer-specific case-control studies were all approved by the ethics committee of IARC and participating EPIC centers.

**Table 1.** Description of the original seven cancer-specific matched case-control studies nested within EPIC

| Cancer site | Number of Samples | Matrix | Laboratory | Kit Used |
| --- | --- | --- | --- | --- |
| Breast | 3,172 | Citrate plasma<sup>1</sup> | IARC | p180 |
| Colorectal (Study 1) | 946 | Citrate plasma | IARC | p180 |
| Colorectal (Study 2) | 2,295 | Serum | HZM<sup>2</sup> | p150 |
| Endometrial | 1,706 | Citrate plasma | ICL<sup>3</sup> | p180 |
| Liver | 662 | Serum | IARC | p180 |
| Kidney | 1,213 | Citrate plasma | IARC | p180 |
| Prostate | 6,020 | Citrate plasma | IARC | p180 |

<sup>1</sup>except Swedish participants (n=101; EDTA plasma). <sup>2</sup>Helmhotz Zentrum München. <sup>3</sup>Imperial College London

### Laboratory analysis

As summarized in Table 1, pre-diagnostic blood samples were assayed at the Helmholtz Zentrum (München, Germany) for the second colorectal cancer study, at Imperial College London (UK) for the endometrial cancer study, and at IARC for all other studies. Data for a total of 171 metabolites were acquired by tandem mass spectrometry using either the AbsoluteIDQ p150 (for the second colorectal cancer study) or the AbsoluteIDQ p180 commercial kit (Biocrates Life Science AG, Innsbruck Austria). Two successive assays were used, liquid chromatography-tandem mass spectrometry (LC-MS/MS) for amino acids and biogenic amines, and flow injection analysis-tandem mass spectrometry (FIA-MS/MS) for the other metabolites. Samples were either serum or citrate plasma, and samples within each study were all from the same type of blood matrix, except for the breast cancer study (Table 1).

### Selection of the metabolites, data pre-processing

Data were pre-processed following an established procedure^30^. Briefly, metabolites with more than 25% missing values in any study were excluded. Samples with more than 25% missing values overall were excluded, as were those detected as outliers by a principal component analysis (PCA)-based approach applied within each study separately. Then, for all metabolites measured by FIA with a semi-quantitative method (acylcarnitines, glycerophospholipids, sphingolipids, hexoses), measurements below the batch-specific limit of detection (LOD) were imputed to half the LOD. When the batch-specific LOD was unknown, LOD was first set to study-specific medians of known batch-specific LODs. For the metabolites measured with a fully quantitative approach (amino acids and biogenic amines), measurements below the lower limit of quantification (LLOQ) or above the upper limit of quantification (ULOQ) were imputed to half the LLOQ or to the ULOQ, respectively. For all metabolites, other missing values were imputed to the batch-specific median of the non-missing measurements. The resulting measurements were then log-transformed to improve symmetry.

### Cancer types and exclusion criteria

We focused on eight cancer types, namely breast, colorectal, endometrial, kidney, gallbladder and biliary tract cancers, HCC, advanced and localized prostate. As detailed in Section 1 of the Supplementary Material (Additional file 1), matched case-control pairs for HCC and gallbladder and biliary tract cancer were extracted from the liver cancer study, while matched case-control pairs for advanced and localized prostate cancer were extracted from the prostate cancer study. Since hormones could affect metabolite levels and their association with cancer risk^11^, women using exogenous hormones (either hormone replacement therapy or oral contraceptive) at baseline were excluded.

### Statistical analyses

All analyses were performed using R software. Characteristics of cases and controls for the eight studied cancer types were described using mean and standard deviation or frequency. Pearson correlations between the metabolites were computed in controls only to reduce collider bias.

### Clustering of metabolites

The most strongly correlated metabolites were grouped together by applying the hierarchical clustering approach implemented in the ClustOfVar R package^31^ to the control samples. For each cluster, the method defined its representative as the first principal component in the PCA of the metabolites grouped into that cluster. In our figures and tables, cluster representatives were labeled as “xxx_clus*”*, with “xxx*”* representing one particular metabolite that composed that cluster. We retained the model with lowest number of clusters such that representatives explained at least 80% of the total variation in each cluster. Cluster representatives and metabolites left isolated after the clustering were simply referred to as metabolites hereafter.

### Multivariate analyses

Given the number of studied metabolites, penalized conditional logistic regression models were used to estimate mutually adjusted associations with cancer risk. Since body-mass index (BMI) could be a strong confounder of the relationship between several of the examined metabolites^32,33^ and cancers^34–38^, metabolite-specific linear models were used to compute residuals on BMI. To account for the large number of metabolites and leverage possible commonalities among the metabolic disorders preceding cancer development for different cancer types, estimation was based on the data shared lasso^26–28^, an extension of the lasso^39^ allowing the analysis of case-control studies with multiple disease types. For each metabolite, the data shared lasso decomposes its type-specific odds-ratio as the product of *(i)* an overall odds-ratio capturing the overall association with cancer, and *(ii)* type-specific deviations from this overall odds-ratio. Then, the method identifies whether its overall (mutually adjusted) association with cancer is null or not, and also whether some of its type-specific associations deviate from its (possibly null) overall association with cancer. Compared to more standard approaches, the data shared lasso was shown to perform particularly well for the identification of features with a consistent non-null association with multiple disease types, while also allowing for the identification of type-specific associations^28^.

To assess the robustness of the identified associations, the data shared lasso was applied repeatedly on 100 bootstrap samples generated from the original sample^40^. Moreover, following the rationale of the lasso-OLS hybrid^41^, associations identified by the data shared lasso were further inspected using unpenalized conditional logistic regression models, *(i)* to quantify their strength and investigate possible heterogeneity among the type-specific associations beyond those identified by the data shared lasso (see Section 3 in Additional file 1 for details); *(ii)* to assess possible departure from linearity by comparing models with natural cubic splines to models with linear terms only; and *(iii)* to assess possible attenuation after excluding, in turn, first two and first seven years of follow-up (to examine potential reverse causation and more generally assess the impact of time to diagnosis on our findings), and after adjustment for additional factors (education level, waist circumference, height, physical activity, smoking status, alcohol intake, use of non-steroidal anti-inflammatory drugs, and, for women, menopausal status and phase of menstrual cycle in premenopausal women). Finally, effect modification by BMI was assessed under standard (i.e., non-conditional) logistic regression models after breaking the matching and correcting metabolite measurements for batch and study effects^30^.

### Univariate analyses

For comparison, non-mutually adjusted associations with cancer risk were estimated for each metabolite in conditional logistic regression models adjusted for BMI. Each cancer type was first modelled separately, and then jointly, via one global conditional logistic regression model. Heterogeneity of associations across cancer types was tested by comparing the difference in log-likelihood between the global model and a model with interaction terms between each metabolite and cancer type to a chi-square distribution with 8-1=7 degrees of freedom. To account for multiple comparisons, associations and heterogeneities with a False Discovery Rate (FDR) inferior to 5% were considered as statistically significant^42^.

### Analysis of additional metabolites

The 16 metabolites (Table S1, Additional file 2) that were not acquired in the second colorectal cancer study (AbsoluteIDQ p150 kit) were not included in our main analysis and were examined in a reduced sample, using the methods described above.

## Results

### Description of the study population

After the exclusions of subjects detailed in Figure 1, 11,656 EPIC participants were included in the analysis comprising 5,828 matched case-control pairs. Cases were diagnosed at an average age of 64.4 years, 8.4 years after blood collection. The main characteristics of cases and controls in each study are displayed in Table 2. The main analysis focused on 117 metabolites that were retained after the pre-processing step (Table S1, Additional file 2). As displayed in Figure S1 in Additional file 2, strong positive correlations were observed between some metabolites, particularly between some of the glycerophospholipids (phosphatidylcholines, PCs, and lysophosphatidylcholines, lysoPCs), and sphingomyelins (SMs).

**Table 2.** Main characteristics of the control (Ctrl) and case (Case) sub-populations in the eight cancer type-specific EPIC studies

|  | BrC study |  | CRC study |  | EnC study |  | KiC study |  | GBTC study |  | HCC study |  | Adv. PrC study |  | Loc. PrC study |  |
| --- | --- | --- | --- | --- | --- | --- | --- | --- | --- | --- | --- | --- | --- | --- | --- | --- |
|  | N = 1,088 pairs |  | N = 1,500 pairs |  | N = 689 pairs |  | N = 511 pairs |  | N = 85 pairs |  | N = 121 pairs |  | N = 533 pairs |  | N = 1301 pairs |  |
|  | Ctrl | Case | Ctrl | Case | Ctrl | Case | Ctrl | Case | Ctrl | Case | Ctrl | Case | Ctrl | Case | Ctrl | Case |
| **Age at blood collection** |  |  |  |  |  |  |  |  |  |  |  |  |  |  |  |  |
| Mean | 51.8 | 51.8 | 57.0 | 57.1 | 54.3 | 54.3 | 55.8 | 55.8 | 58.7 | 58.7 | 59.9 | 59.9 | 57.6 | 57.6 | 57.9 | 58.0 |
| (SD) | (8.31) | (8.33) | (7.58) | (7.57) | (7.83) | (7.84) | (8.47) | (8.46) | (7.13) | (7.08) | (7.01) | (6.98) | (7.18) | (7.18) | (6.80) | (6.80) |
| **Age at cancer diagnosis** |  |  |  |  |  |  |  |  |  |  |  |  |  |  |  |  |
| Mean | - | 60.4 | - | 64.9 | - | 62.7 | - | 64.5 | - | 64.9 | - | 66.1 | - | 66.3 | - | 67.1 |
| (SD) | - | (8.83) | - | (8.18) | - | (8.16) | - | (8.83) | - | (7.60) | - | (7.49) | - | (7.02) | - | (6.36) |
| **Sex** |  |  |  |  |  |  |  |  |  |  |  |  |  |  |  |  |
| Female | 1088 | 1088 | 769 | 769 | 689 | 689 | 197 | 197 | 48 | 48 | 35 | 35 | 0 (0%) | 0 (0%) | 0 (0%) | 0 (0%) |
|  | (100%) | (100%) | (51.3%) | (51.3%) | (100%) | (100%) | (38.6%) | (38.6%) | (56.5%) | (56.5%) | (28.9%) | (28.9%) |  |  |  |  |
| **BMI (kg/m<sup>2</sup>)** |  |  |  |  |  |  |  |  |  |  |  |  |  |  |  |  |
| Mean | 25.7 | 26.2 | 26.5 | 27.2 | 26.0 | 28.2 | 26.7 | 27.8 | 26.9 | 27.3 | 26.9 | 28.4 | 26.7 | 27.0 | 27.5 | 27.2 |
| (SD) | (4.32) | (4.80) | (3.88) | (4.34) | (4.29) | (5.52) | (3.84) | (4.47) | (4.38) | (3.98) | (3.72) | (4.73) | (3.48) | (3.20) | (3.49) | (3.37) |
| **Education** |  |  |  |  |  |  |  |  |  |  |  |  |  |  |  |  |
| None | 56 | 62 | 124 | 136 | 67 | 76 | 42 | 34 | 6 | 6 | 6 | 7 | 28 | 32 | 106 | 136 |
|  | (5.1%) | (5.7%) | (8.3%) | (9.1%) | (9.7%) | (11.0%) | (8.2%) | (6.7%) | (7.1%) | (7.1%) | (5.0%) | (5.8%) | (5.3%) | (6.0%) | (8.1%) | (10.5%) |
| Primary school completed | 397 | 377 | 574 | 526 | 269 | 231 | 184 | 186 | 33 | 39 | 49 | 52 | 160 | 166 | 444 | 411 |
|  | (36.5%) | (34.7%) | (38.3%) | (35.1%) | (39.0%) | (33.5%) | (36.0%) | (36.4%) | (38.8%) | (45.9%) | (40.5%) | (43.0%) | (30.0%) | (31.1%) | (34.1%) | (31.6%) |
| Technical/professional school | 245 | 254 | 333 | 334 | 124 | 118 | 107 | 109 | 20 | 15 | 31 | 38 | 140 | 127 | 305 | 306 |
|  | (22.5%) | (23.3%) | (22.2%) | (22.3%) | (18.0%) | (17.1%) | (20.9%) | (21.3%) | (23.5%) | (17.6%) | (25.6%) | (31.4%) | (26.3%) | (23.8%) | (23.4%) | (23.5%) |
| Secondary school | 158 | 178 | 188 | 227 | 100 | 127 | 66 | 72 | 11 | 8 | 11 | 5 | 58 | 59 | 103 | 91 |
|  | (14.5%) | (16.4%) | (12.5%) | (15.1%) | (14.5%) | (18.4%) | (12.9%) | (14.1%) | (12.9%) | (9.4%) | (9.1%) | (4.1%) | (10.9%) | (11.1%) | (7.9%) | (7.0%) |
| Longer education (incl. University deg.) | 211 | 195 | 241 | 227 | 100 | 100 | 96 | 93 | 15 | 17 | 22 | 17 | 132 | 124 | 306 | 324 |
|  | (19.4%) | (17.9%) | (16.1%) | (15.1%) | (14.5%) | (14.5%) | (18.8%) | (18.2%) | (17.6%) | (20.0%) | (18.2%) | (14.0%) | (24.8%) | (23.3%) | (23.5%) | (24.9%) |
| Not specified | 21 | 22 | 40 | 50 | 29 | 37 | 16 | 17 | 0 | 0 | 2 | 2 | 15 | 25 | 37 | 33 |
|  | (1.9%) | (2.0%) | (2.7%) | (3.3%) | (4.2%) | (5.4%) | (3.1%) | (3.3%) | (0%) | (0%) | (1.7%) | (1.7%) | (2.8%) | (4.7%) | (2.8%) | (2.5%) |

**Figure 1.**
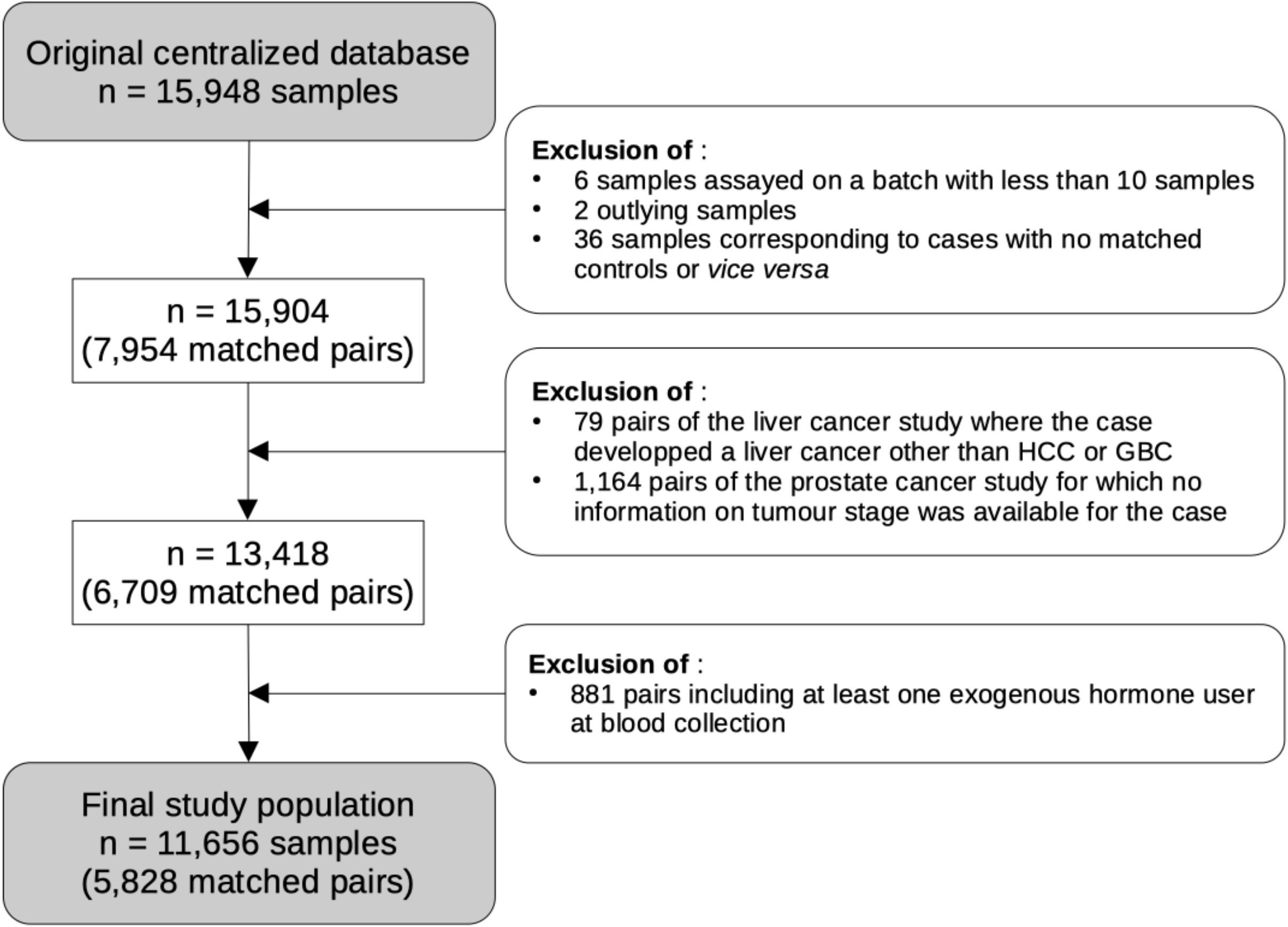
Flowchart summarizing the exclusion criteria to derive the final sample used in our main analysis. GBC stands for gallbladder and biliary tract cancer and HCC for hepatocellular carcinoma.

### Clustering of metabolites

The hierarchical clustering applied to controls grouped 100 metabolites into 33 clusters of size ranging from 2 to 6 metabolites per cluster, while 17 metabolites remained isolated. As displayed in Figure 2, clusters comprised metabolites of the same chemical class, and correlations between metabolites and their representative were consistently greater than 0.83. On average, clusters’ representatives explained 86% of the total variation of their cluster (range: 80%-95%), and the 33 + 17 = 50 studied metabolites together explained more than 88% of the total variation of the original 117 metabolites.

**Figure 2.**
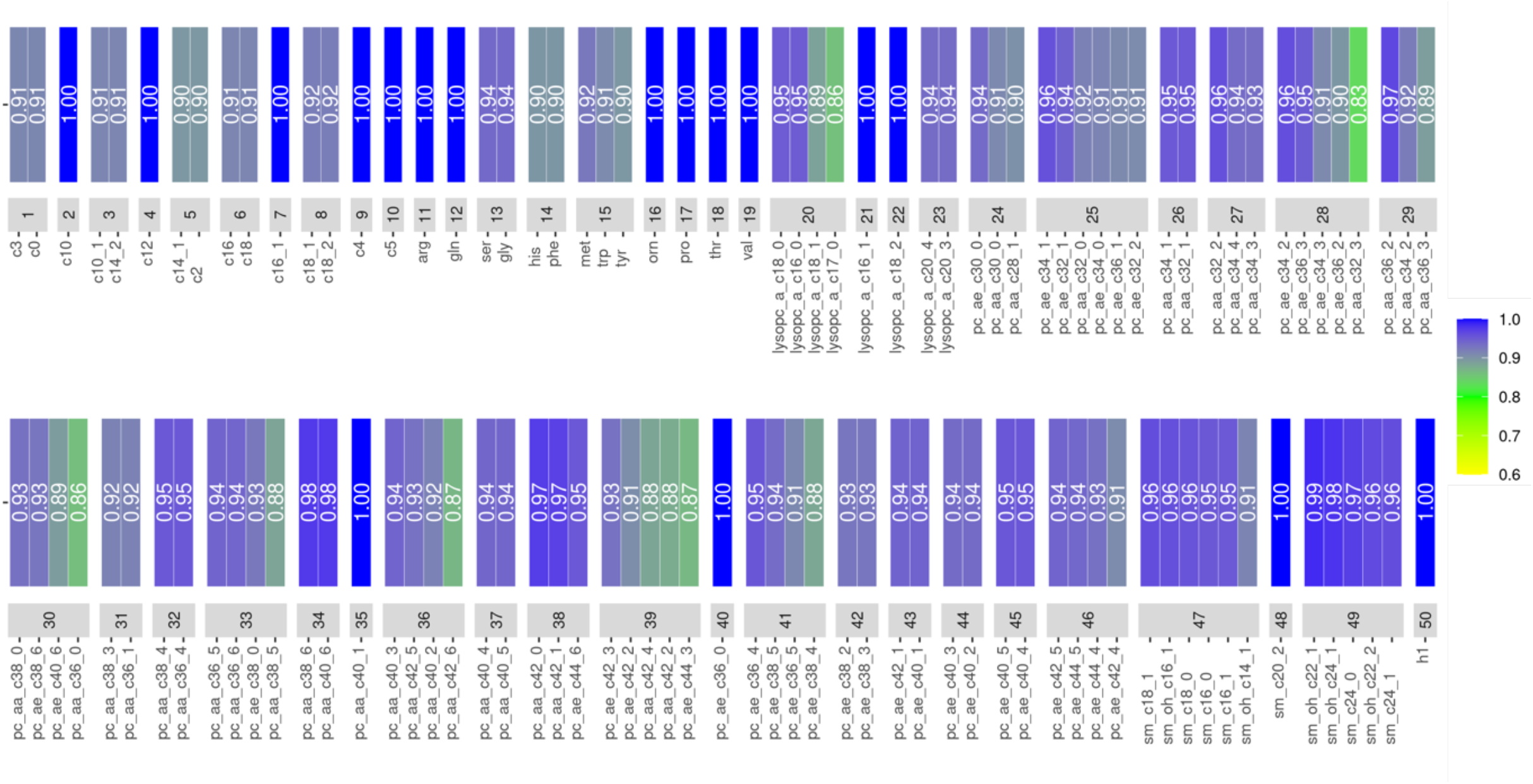
Description of the 50 “metabolites” retained for the main analysis, which includes 33 clusters of strongly correlated metabolites and 17 “isolated” metabolites. For example, the 19^th^ metabolite is an isolated metabolite (valine), while the 26^th^ one is a cluster made of two phosphatidylcholines. For each cluster, correlations between its representative and the individual metabolites that compose that cluster are represented as a heat map (this correlation is 1 when the “cluster” is actually reduced to an isolated metabolite)

### Multivariate analyses

As displayed in Figures 3 and 4, the data shared lasso identified nine metabolites with a non-null overall association with cancer: butyrylcarnitine (acylcarnitine C4), glutamine, lysoPC a C18:2, and three clusters of PCs (those containing PC aa C32:2, PC aa C36:0, and PC aa C36:1, respectively), with an inverse overall association with cancer risk, and decanoylcarnitine (acylcarnitine C10), proline and the cluster of PCs that included PC aa C28:1 with a positive overall association. Cancer type-specific deviations from the overall association with cancer risk were identified for three of these metabolites: the association between proline and breast cancer risk was inverse or null, while the associations between lysoPC a C18:2 and the cluster containing PC aa C36:0 with localized prostate cancer were positive or null.

**Figure 3.**
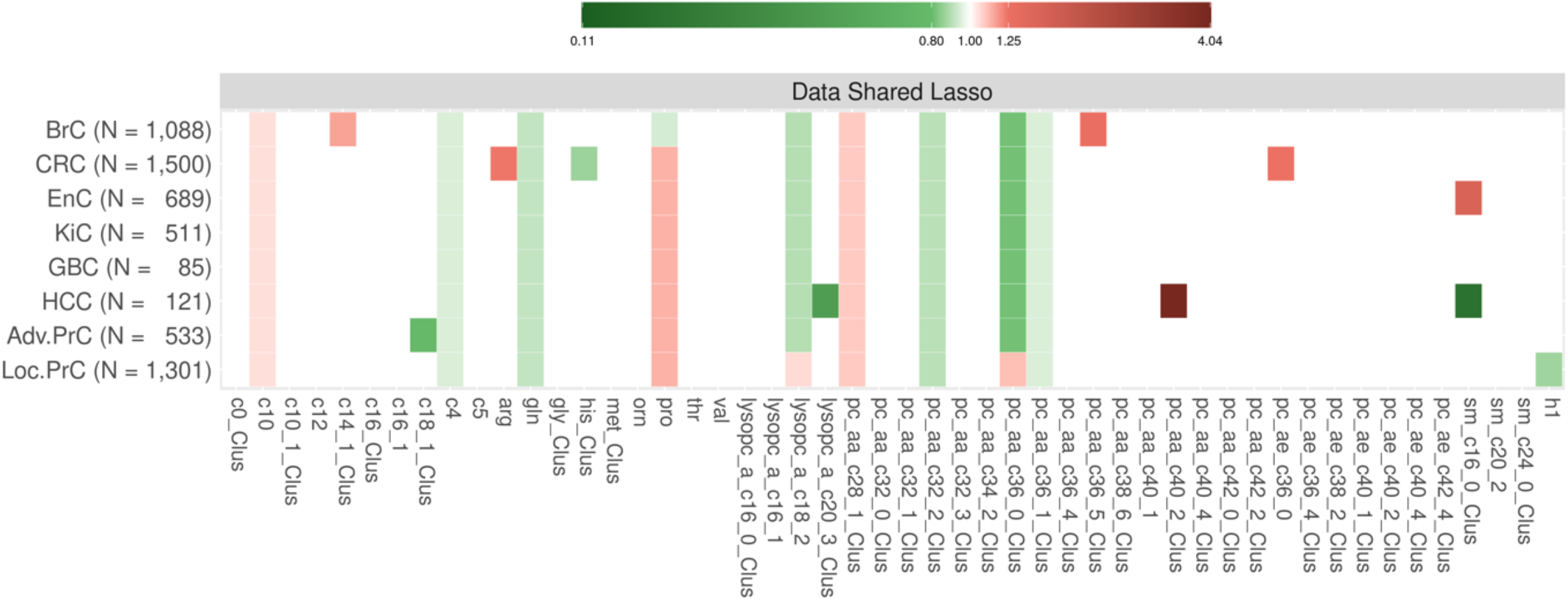
Summary of the main results from our the multivariate pan-cancer analysis, which evaluated mutually adjusted associations between each feature (more precisely, their residuals after adjustment for BMI) and the risk of the eight cancer types, using a data shared lasso penalty. White entries correspond to the absence of identified associations, while green and red entries correspond to inverse and positive associations, respectively. The more intense the colour, the larger the absolute value of the log-odds-ratio (that were re-estimated in multivariate unpenalized conditional regression models; see Section 3.a in the Supplementary Material for details). The x-axis represents the 50 features (33 cluster representatives and 17 isolated metabolites). In the labels of the y-axis, numbers correspond to numbers of pairs for each type-specific cancer (and in total for the pooled analysis), while BrC stands for breast cancer, CRC for colorectal cancer, EnC for endometrial cancer, KiC for Kidney cancer, GBC for gallbladder and biliary tract cancer, HCC for hepatocellular carcinoma, and Adv.PrC and Loc.PrC for advanced and localized prostate cancers, respectively.

**Figure 4.**
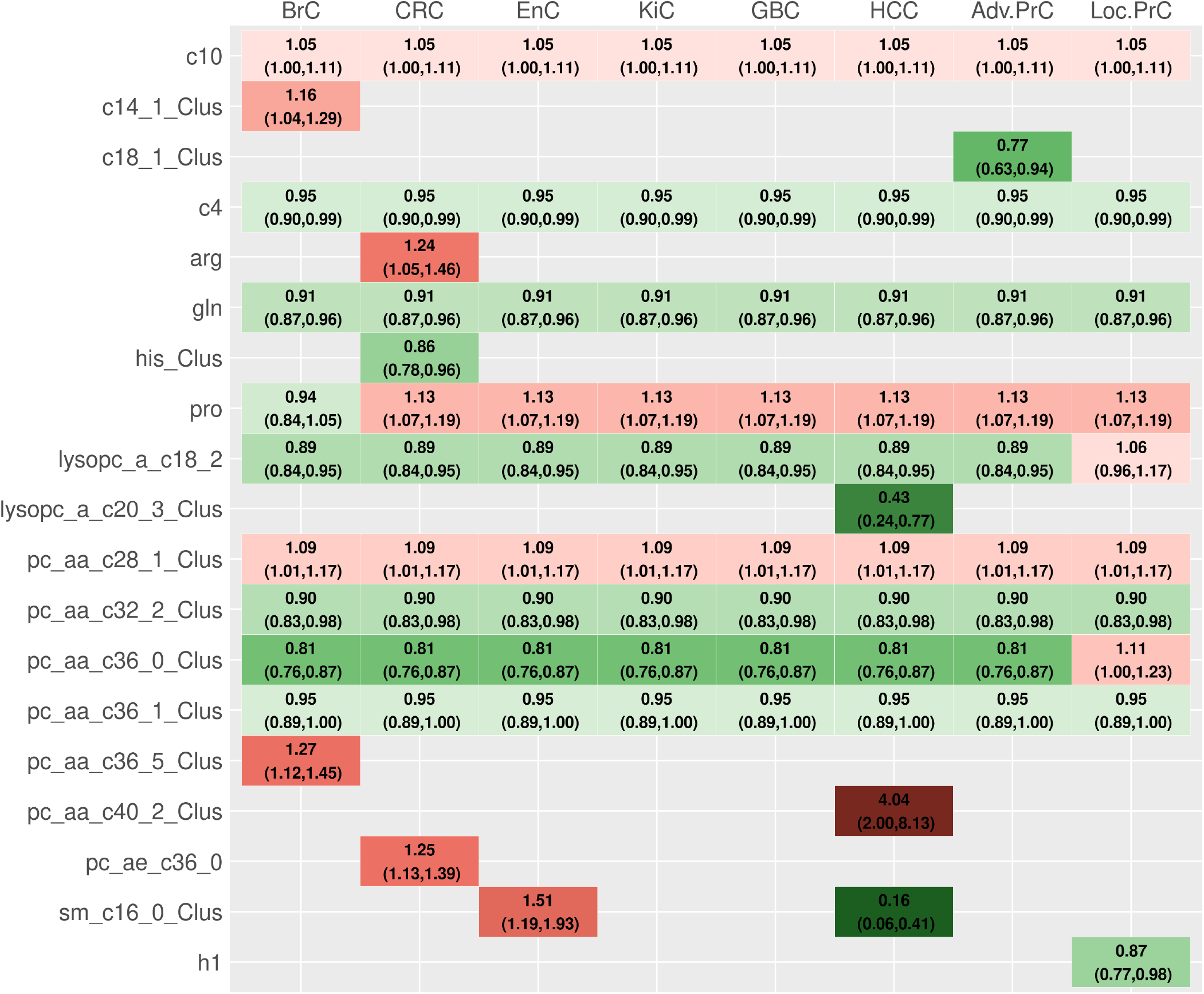
Summary of the mutually adjusted associations between the 50 metabolic features and risks of the eight cancer types, as identified by the data shared lasso. Only the 19 features (8 isolated metabolites and 11 cluster representatives) for which the data shared lasso identified an association with at least one cancer type are presented on the *y* axis. Point estimates and 95% confidence intervals of the corresponding odds-ratios were obtained through non-penalized conditional logistic regression models using the design matrix derived from the positions of the non-zero components in the data shared lasso vector estimate 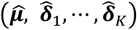; see Section 3.a in the Supplementary Material for details. They have to be interpreted with caution since they are the result of post-selection inference. In the labels of the columns, BrC stands for breast cancer, CRC for colorectal cancer, EnC for endometrial cancer, KiC for Kidney cancer, GBC for gallbladder and biliary tract cancer, HCC for hepatocellular carcinoma, and Adv.PrC and Loc.PrC for advanced and localized prostate cancers, respectively.

Several cancer type-specific associations were identified among the remaining 41 metabolites. Specifically, positive associations were observed between breast cancer risk and two clusters, that included tetradecenoylcarnitine (acylcarnitine C14:1) and PC aa C36:5, respectively. Risk of colorectal cancer was positively associated with arginine and PC ae C36:0, and inversely associated with the cluster that included histidine. Risk of HCC was positively associated with the cluster containing PC aa C40:2, and inversely associated with the two clusters that included lysoPC a C20:3 and SM C16:0, respectively. This latter cluster was also positively associated with endometrial cancer risk. The cluster that included octadecenoylcarnitine (acylcarnitine C18:1) was inversely associated with risk of advanced prostate cancer. Finally, risk of localized prostate cancer was inversely associated with hexoses (H1).

The strength of the associations identified by the data shared lasso was similar after excluding, in turn, the first two and the first seven years of follow-up (Figure S2, Additional file 2). Likewise, models adjusted for additional factors produced similar associations (Figure S2, Additional file 2), except for the overall association with cancer for the cluster that included PC aa C28:1, whose odds-ratio (OR) was attenuated from 1.09 (95% confidence interval: 1.01-1.17) to 1.04 (0.98-1.12), and for the association between endometrial cancer risk and the cluster that included SM C16:0, whose OR decreased from 1.51 (1.19-1.93) to 1.20 (0.97-1.47). For each overall association and type-specific deviation identified by the data shared lasso, linearity and absence of effect modification by BMI were compatible with our data (Figure S3, Additional file 2). Focusing on the nine metabolites that had a non-null overall association with cancer, the analysis presented in Figure S4 in Additional file 2 suggested possible cancer type-specific deviations from the overall associations beyond the three ones identified by the data shared lasso, in particular for HCC (with acylcarnitine C4, proline and the cluster that comprises PC aa C36:1) and for kidney cancer (with acylcarnitines C10 and C4 and the cluster that comprises PC aa C36:1). However, none of the comparisons between the models identified by the data shared lasso and the nine “extended” models used to derive these fully cancer-type specific associations reached statistical significance (Figure S4, Additional file 2).

As displayed in Table 3 (third column), 15 out of the 22 associations identified by data shared lasso were replicated in more than 50% of the bootstrap samples. As displayed in Table 4, three inverse cancer type-specific associations that were not identified by the data shared lasso on the original sample were identified in more than 55% of the bootstrap samples: the cluster comprising glycine with endometrial cancer risk (identified in 65% of the bootstrap samples), the cluster containing decenoylcarnitine (acylcarnitine C10:1) with risk of kidney cancer (56%) and lysoPC a C16:1 with risk of localized prostate cancer (84%). Positive associations between arginine and kidney cancer risk (74%) and between the cluster containing lysoPC a C16:0 and localized prostate cancer risk (86%) were also observed in more than 55% of the bootstrap samples.

**Table 3:** Robustness of the associations identified in the main analysis. For each identified association, the proportion of bootstrap samples on which it was replicated is reported (in bold when ≥50%).

| Feature | Cancer Type* | Proportion of bootstrap samples<sup>1</sup> | Proportion of bootstrap samples<sup>2</sup> |
| --- | --- | --- | --- |
| **Overall associations with cancer risk** |  |  |  |
| c10 | Overall | **62%** | **59%** |
| c4 | Overall | 47% | 39% |
| gln | Overall | **73%** | **76%** |
| pro | Overall | **65%** | **50%** |
| lysopc_a_c18_2 | Overall | **57%** | 47% |
| pc_aa_c28_1_Clus | Overall | **57%** | **64%** |
| pc_aa_c32_2_Clus | Overall | 49% | **71%** |
| pc_aa_c36_0_Clus | Overall | **86%** | **95%** |
| pc_aa_c36_1_Clus | Overall | **50%** | 40% |
| **Cancer type-specific associations** |  |  |  |
| c14_1_Clus | BrC | **80%** | **76%** |
| pro | BrC | **77%** | **70%** |
| pc_aa_c36_5_Clus | BrC | 36% | 47% |
| arg | CRC | **88%** | 19% |
| his_Clus | CRC | **81%** | **72%** |
| pc_ae_c36_0 | CRC | **80%** | 46% |
| sm_c16_0_Clus | EnC | **85%** | **87%** |
| lysopc_a_c20_3_Clus | HCC | 32% | 47% |
| pc_aa_c40_2_Clus | HCC | **61%** | 34% |
| sm_c16_0_Clus | HCC | **90%** | **78%** |
| c18_1_Clus | Adv.PrC | 40% | 49% |
| lysopc_a_c18_2 | Loc.PrC | 14% | 23% |
| pc_aa_c36_0_Clus | Loc.PrC | 49% | 41% |

\* BrC stands for breast cancer, CRC for colorectal cancer, EnC for endometrial cancer, HCC for hepatocellular carcinoma, and Adv.PrC Loc.PrC for advanced and localized prostate cancers, respectively.
<sup>1</sup> Bootstrap samples were generated from the original sample of 5,828 matched case-control pairs with information on 117 metabolites (corresponding to 50 features after the clustering step).
<sup>2</sup> Bootstrap samples were generated from the original sample which comprised 4,761 matched case-control pairs with information on 133 metabolites (corresponding to 65 features after the clustering step) after excluding the participants of the second CRC study.

**Table 4:**
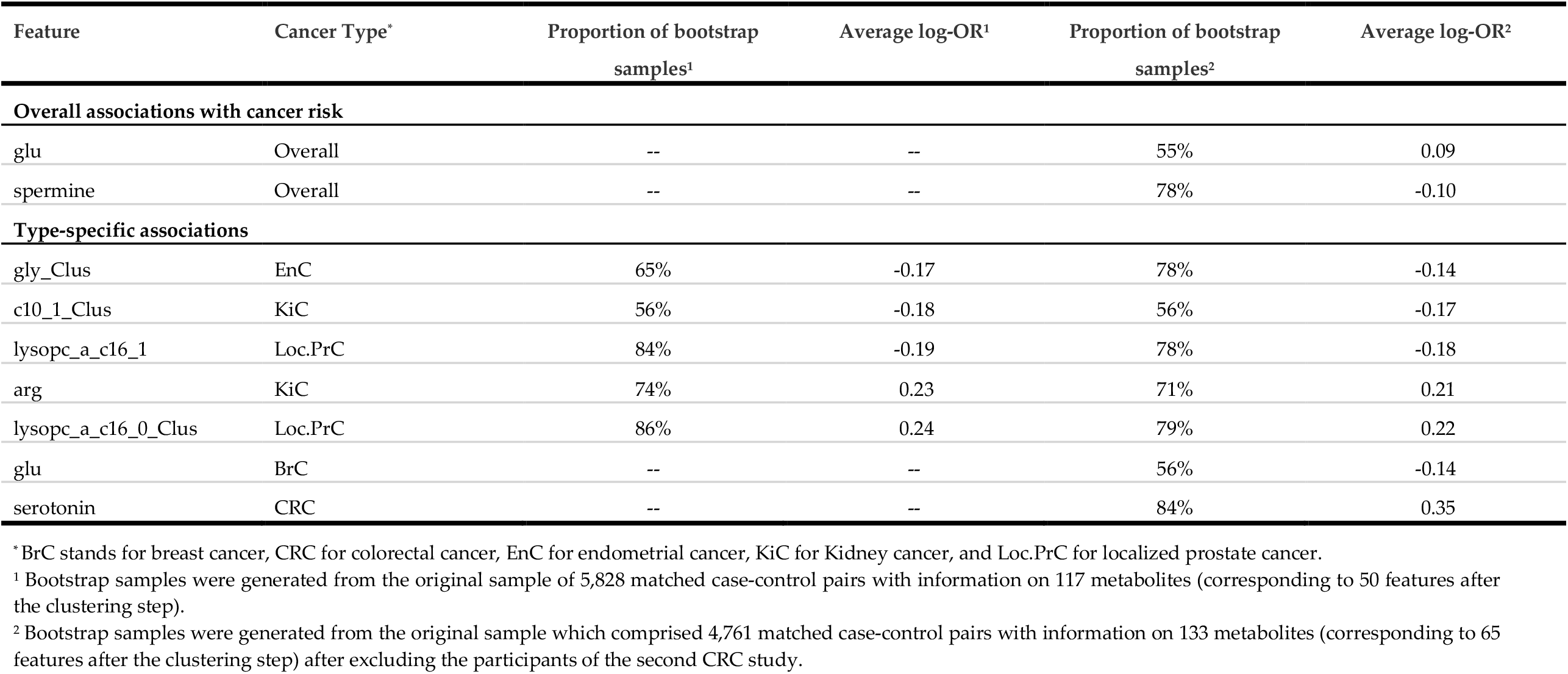
Other associations identified in a large proportion of the bootstrap samples. Associations identified in at least 55% of both bootstrap analyses are reported, along with the proportion of bootstrap samples in which they were identified, and the corresponding average log odds-ratio (as estimated by the data shared lasso on each bootstrap sample).

### Analysis of the extended list of metabolites

After excluding 2,134 samples from the second colorectal cancer study which used a different platform that measured a lower number of metabolites, 16 additional metabolites could be evaluated (Table S1, Additional file 2). Among them, the clustering step grouped leucine and isoleucine together. The analysis of this extended list of metabolites then focused on 65 metabolites (31 isolated metabolites and 34 cluster representatives), measured in 9,522 participants. As displayed in Table 3, 11 out of the 22 associations identified in the main analysis presented above were again replicated in more than 50% of the bootstrap samples generated from this reduced sample. Four associations that were not identified in our previous analyses were identified in more than 55% of these new bootstrap samples (Table 4): an overall positive association between cancer risk and glutamate (55% of the bootstrap samples), an overall inverse association between cancer risk with spermine (78%), as well as two cancer type-specific associations between glutamate with breast cancer risk (inverse, 56%) and between serotonin and colorectal cancer risk (positive, 84%).

### Univariate analyses

As displayed in Figure S5 in Additional file 2, the cancer type-specific univariate analyses identified associations with risks of breast cancer (two positive associations and four inverse), colorectal cancer (three inverse), endometrial cancer (two inverse), kidney cancer (one inverse), HCC (four positive associations and 16 inverse) and advanced prostate cancer (seven inverse associations). The univariate pooled analysis identified 15 inverse associations, and there was no evidence of heterogeneity across cancer type for four of them (butyrylcarnitine, and the three clusters containing PC aa C32:2, PC ae C36:4, and PC ae C38:2, respectively).

## Discussion

Using available metabolomics data from eight cancer-specific matched case-control studies nested within the EPIC cohort, we investigated the relationship between pre-diagnostic blood levels of over one hundred metabolites and risks of breast cancer, colorectal cancer, endometrial cancer, gallbladder and biliary tract cancer, HCC, kidney cancer, and localized and advanced prostate cancers. In our main analysis, we found nine metabolites associated with cancer risk across different cancer types, suggesting the existence of shared metabolic pathways, as well as fourteen cancer-type specific associations. These identified associations were found to be robust after extensive sensitivity analyses: in particular, they were not attenuated after exclusion of the first years of follow-up, hence were less likely to be due to reverse causality, were not attenuated after adjustment for relevant cancer risk factors, were not modified by BMI, and did not deviate significantly from linearity. In additional analyses, in particular those based on bootstrap samples, we identified several additional metabolites possibly associated with risk of specific cancer types or with cancer risk across different cancer types.

Our results suggested that concentrations of glycerophospholipids (phosphatidylcholines and lysophosphatidylcholines) could be linked to the risk of cancer overall as well as to specific cancer types. The role of glycerophospholipids in carcinogenesis is not fully understood but could be related to their documented anti-inflammatory properties, protection from oxidative stress, inhibition of cell proliferation and induction of apoptosis^43–45^. We observed a consistent inverse association between cancer risk with lysoPC a C18:2 as well as three clusters of phosphatidylcholines across all studied cancer types, except localized prostate cancer for which the association with lysoPC a C18:2 and one cluster of phosphatidylcholines was absent, or positive. An inverse association was previously reported between lysoPC a C18:2 with T2D in different studies^7,46^ as well as with risks of breast, colorectal and prostate cancers in the pan-cancer analysis conducted in the EPIC Heidelberg study^25^. Our results regarding the three clusters of phosphatidylcholines were in line with many previously reported inverse associations between cancer and phosphatidylcholines^11,12,15,16,20,47^. Besides, we identified a positive association between the cluster that included PC aa C28:1 and cancer risk across all studied cancer types. This cluster also comprised PC ae C30:0, for which a positive association was reported with risks of breast, colorectal and prostate cancers in the EPIC Heidelberg study^25^. Cancer type-specific positive associations were found for the cluster containing PC aa C36:5 with breast cancer, PC ae C36:0 with colorectal cancer, and the cluster containing PC aa C40:2 with HCC. These three clusters were correlated with one another (Pearson correlation greater than 0.48), indicating that higher levels of these phosphatidylcholines might contribute to the development of these three cancer types.

We also observed robust associations between specific circulating amino acids and cancer risk. Our results suggested that proline was positively related to cancer risk across all studied cancer types, except breast cancer and possibly HCC (see Figure S4 in Additional file 2). A positive association between proline and prostate cancer risk was previously reported in EPIC^12^. In addition, a drosophila model of high-sugar diet^48^ recently highlighted the possible role of proline in tumour growth, and proline was also found to distinguish colorectal cancer patients from those with adenomas^49^, and to be associated with metastasis formation^50^. Glutamine was inversely associated with overall cancer risk in our analysis, while glutamate, a metabolite of glutamine, was positively related to the risk of all cancer types except for breast cancer. Although prior studies of the French E3N and SU.VI.MAX cohorts reported a positive association between glutamine and premenopausal breast cancer^51,52^, our results regarding glutamine and glutamate were consistent with those of many previous studies that reported inverse associations between glutamine and risk of colorectal cancer^18^, HCC^19,53^ and T2D^7,54^, and positive associations between glutamate and risk of premenopausal breast cancer^52^, kidney cancer^15^, HCC^19,53^, as well as T2D^7^. Lower serum levels of glutamine were also observed in kidney cancer^55^ and ovarian cancer^56^ cases compared to controls. Glutamine is an energy substrate for cancer cells and makes a major contribution to nitrogen metabolism. Alterations in glutamine-glutamate equilibrium often reflect energetic processes related to cancer metabolism^57^. It is possible that altered levels of glutamine and glutamate in individuals subsequently diagnosed with cancer may reflect ongoing metabolic processes related to cancer development and as such may serve as an early biomarker of cancer risk. However, the inverse association between glutamine levels and overall cancer risk observed in our analysis was only slightly attenuated after excluding, in turn, the first two and the first seven years of follow-up suggesting that changes in the glutamine-glutamate may precede cancer development.

Our analysis additionally identified two positive and two inverse cancer type-specific associations with circulating amino acids. We observed an inverse association between colorectal risk and the cluster containing histidine, for which previous studies reported inverse associations with risks of colorectal cancer and T2D^54^, while a positive association was reported with breast cancer^52^. Also, lower serum levels of histidine were previously reported in ovarian cancer cases compared to controls^58^. Our results further suggested an inverse association between endometrial cancer risk and the cluster composed of glycine and serine, in line with previous results from the EPIC cancer-specific study of endometrial cancer^14^. Previous studies also reported inverse associations between glycine and/or serine with risks of T2D^54^. Finally, our analysis suggested a positive association between arginine with risks of colorectal and kidney cancers (Table 4). Arginine was previously found to be positively associated with breast cancer in the E3N cohort^52^, while an inverse association with breast cancer was reported in EPIC^11^.

Regarding the biogenic amines, we found a positive association between serotonin levels and colorectal cancer risk, consistent with previous results from the CORSA case-control study and a previous EPIC analysis of colon cancer^59^. We also found a consistent inverse association between spermine and risk of the eight studied cancer types. Like other polyamines, spermine is involved in cell proliferation and differentiation and has antioxidant properties^60^, and dysregulation of polyamines metabolism is characteristic of multiple types of tumours^61^. It was previously reported that polyamine supplementation, in particular spermidine, which acts as an intermediate in the conversion of putrescine to spermine, could be related to reduced overall and cancer-specific mortality^62–64^.

In our analysis, localized and advanced prostate cancers were considered as two different outcomes as previous results suggested that metabolic dysregulation might be predictive of advanced or aggressive prostate cancers only^12^. In fact, we observed some differences between the metabolites associated with risks of localized and advanced prostate cancers, respectively. Specifically, and as previously reported^12,13^, our results suggested that hexoses, glycerophospholipids, octadecenoylcarnitine and/or octadecadienylcarnitine could help differentiate the respective mechanisms involved in the development of aggressive and localized prostate tumours.

Some metabolites identified in our study were previously associated with established cancer risk factors, such as obesity ^32,33^. In particular, a recent metabolomics study of BMI reported inverse associations with glutamine, lysophosphatidylcholine a C18:2 and phosphatidylcholine PC aa C38:0 (which was clustered with PC aa C36:0 in our analysis), and a positive association with glutamate^33^. Directions of the associations with BMI were consistent with those identified in our study with cancer risk after adjustment for BMI, indicating that these metabolites might be mediators of the obesity-cancer relationship.

Our study has several strengths. First, it relied on a large sample of pre-diagnostic metabolomics data acquired among 5,828 case-control pairs in nested studies on eight cancer types within a large prospective cohort, on average 6.4 years before cases developed cancer. Second, in a context where some metabolites might be predictive of cancer risk for multiple cancer types, the data shared lasso used in our analysis automatically accounted for or ignored cancer types when assessing the association between each metabolic feature with cancer risk, depending on whether heterogeneity among the cancer type-specific associations was supported by the data for that particular feature. The comparison of results produced by the standard univariate analyses and the data shared lasso illustrated the interest of the latter. First, the data shared lasso benefited from the increased statistical power of the pooled analysis for the identification of metabolites that could be involved in cancer development for multiple cancer types: for example, butyrylcarnitine (acylcarnitine C4) was not associated with cancer risk in any of the cancer type-specific univariate analyses, while it was in the univariate pooled analysis and in the data shared lasso analysis. Moreover, unlike the simple pooled analysis, the data shared lasso would not necessarily mask cancer type-specific associations: for example, the data shared lasso identified a positive association between the cluster containing tetradecenoylcarnitine (acylcarnitine C14:1) and breast cancer risk, as the univariate analysis of the breast cancer study did, while the univariate pooled analysis could not. Another key difference between the standard univariate analyses and the data shared lasso is that the latter allowed the investigation of mutually adjusted associations, hence the identification of metabolites or clusters of metabolites whose association with cancer risk could not be explained away by other metabolites included in our analysis. Further, mutual adjustment revealed associations that could not be detected in minimally-adjusted models, such as the one between arginine and colorectal cancer risk, which was not apparent in models not adjusted for glutamine and histidine. Another strength of our study stemmed from the extensive sensitivity analyses that we carried out.

On the other hand, identifying cancer risk factors is particularly challenging when candidate risk factors are strongly correlated with one another. Here, we clustered the most strongly correlated metabolites together prior to applying the data shared lasso. As a sensitivity analysis, the data shared lasso was applied to the original set of 117 metabolites, thus ignoring the clustering step, and results were largely consistent with those of our main analysis (Figure S6, Additional file 2). Moreover, because strong correlations remained among some of the metabolites produced by the hierarchical clustering (Figure S7, Additional file 2), we applied the data shared lasso to multiple bootstrap samples to gauge the robustness and specificity of the associations identified in our main analysis. Although most of the identified associations were replicated in a large proportion of bootstrap samples, a few of them were less robust, hence more questionable. For example, the identified inverse association between HCC risk and the cluster that included lysoPC a C20:3 was replicated in 32% of the bootstrap samples only. This lack of robustness could be due to the strong correlation between this cluster and the other three studied metabolites related to lysoPCs (Pearson correlation greater than 0.65; Figure S7 in Additional file 2). As a matter of fact, an inverse association between HCC risk and at least one of the four metabolites related to lysoPCs was identified in 78% of the bootstrap samples. Overall, these results were suggestive of a stronger inverse association with features related to lysoPCs for HCC compared to the other cancer types, but our analysis failed to unambiguously identify which specific lysoPCs might underlie this stronger inverse association. An additional limitation for interpreting the lipid results is the lack of specificity for lipids measured with the AbsoluteIDQ p180/p150 kits as a result of the FIA method^65,66^. Moreover, the limited sample size for some of the studied cancer types (in particular gallbladder and biliary tract cancer and HCC) was a limitation for the identification of cancer type-specific deviations. In this respect, we complemented our analysis by the inspection of estimates computed under models derived from the one identified by the data shared lasso but that further allowed fully type-specific associations (Figure S4, Additional file 2). Another potential limitation of our study was the lack of repeated measurements, yet previous studies suggested that blood levels of metabolites were relatively stable and that a single measurement might be sufficient to capture medium term exposure^67–69^.

## Conclusions

Our results confirmed the complex link between metabolism and cancer risk and highlighted the potential of metabolomics to identify possible informative markers associated with cancer risk and to gain insights into the biological mechanisms leading to cancer development. Our study indicated that specific metabolite families might be related to the risk of multiple cancer types. Some of these metabolites could reflect biological mechanisms underlying the carcinogenic effects of some established cancer risk factors, including obesity.

## Data Availability

The R scripts developed to implement the analyses will be made available on the GitHub platform, for easy access to all interested scientists. The EPIC data is not publicly available, but access requests can be submitted to the Steering Committee (https://epic.iarc.fr/access/submit_appl_access.php).

## List of abbreviations

Adv.PrC: advanced prostate cancer
BMI: body mass index
BrC: breast cancer
CRC: colorectal cancer
CVD: cardio-vascular diseases
EnC: endometrial cancer
EPIC: European Prospective Investigation into Cancer and nutrition
FIA: flow injection analysis
FDR: false discovery rate
GBC: gallbladder and biliary tract cancer
HCC: hepatocellular carcinoma
HZM: Helmholtz Zentrum
IARC: International Agency for Research on Cancer
ICL: Imperial College London
KiC: kidney cancer
Lasso: least absolute shrinkage and selection operator
LC: liquid chromatography
LLOQ: lower limit of quantification
Loc.PrC: localized prostate cancer
LOD: limit of detection
lysoPC: lysophosphatidylcholine
MS/MS: tandem mass spectrometry
OLS: ordinary least square regression
OR: odds ratio
PC: phosphatidylcholine
PCA: principal component analysis
SM: sphingomyelin
T2D: type 2 diabetes
ULOQ: upper limit of quantification.

## Additional files

### Additional file 1

Supplementary material regarding (i) the definition of cancer cases for HCC, GBC, Adv.PrC and Loc.PrC; (ii) the definition and implementation of the data-shared lasso; and (iii) the models used to derive point estimates and confidence intervals from the model selected by the data-shared lasso. (.docx 42 kb)

### Additional file 2

Supplementary tables and figures. Figure S1: Pearson correlation between the 117 original metabolites. Figure S2: sensitivity analyses of mutually adjusted ORs for the overall associations and cancer type-specific deviations. Figure S3: p-values of tests for departure from linearity and effect modification by BMI. Figure S4: ORs for the overall associations identified by the data-shared lasso with (i) the original model (ii) the extended type-specific model. Figure S5: results from the univariate analyses. Figure S6: Comparison of the associations identified by the data shared lasso when working with the 50 features (as in our main analysis) or with the original 117 metabolites. Figure S7: Pearson correlation between the 50 clusters. Table S1: list of the 117 metabolites studied in the main analysis, and of the 16 additional metabolites studied when excluding the second colorectal study. (.docx 2,698 kb)

## Declarations

### Ethics approval and consent

The EPIC study, and in particular the seven case-control studies nested within EPIC, were conducted according to the Declaration of Helsinki, and approved by the ethics committee at the International Agency for Research on Cancer (IARC): on 10 April 2008 (IEC 08-06) and on 11 February 2016 (IEC 16-06) for the liver cancer study, on 7 April 2014 (IEC 14-07) for the breast cancer study, on 7 April 2014 (IEC 14-08) for the two colorectal cancer studies, on 7 April 2014 (IEC 14-09) for the prostate cancer study, on 25 February 2015 (IEC 15-06) for the kidney cancer study, on 28 April 2016 (IEC 16-20) for the endometrial cancer study. Written informed consent was obtained from all subjects involved in the study.

### Authors contributions

The authors responsibilities were as follows: PF, MJG and VV conceived, designed and supervised the research. MB and VV analyzed the data. MB, PF, MJG and VV were responsible for drafting the manuscript. LD, MJen, MJoh, SR, RCT and MJG conducted and supervised metabolomics analyses. LD, MJen, Mjoh, SR, RCT, MH, TJK, JAS, KO, AT, CK, JAR, NL, GS, RK, VK, MSB, FE, DP, SG, SP, RT, CS, BBdM, KSO, TMS, THN, JRQ, CB, MRB, MDC, EA, MS, JM, LV, MR, DM, KT, AKH, HK, JA, PKR, AS and MJG provided the original data, information on the respective populations, and advice on the study design, analysis and interpretation of the results. All authors read and approved of the final manuscript.

### IARC disclaimer

Where authors are identified as personnel of the International Agency for Research on Cancer/World Health Organization, the authors alone are responsible for the views expressed in this article and they do not necessarily represent the decisions, policy, or views of the International Agency for Research on Cancer/World Health Organization.

