## Additional file 1 for "Pan-cancer analysis of pre-diagnostic blood metabolite concentrations in the European Prospective Investigation into Cancer and Nutrition"

*M. Breeur et al.*

### 1. Definitions of cancer cases for HCC, gallbladder and biliary tract cancers (GBC), advanced prostate cancer and localized prostate cancer.

Matched case-control pairs of the liver cancer study were split according to liver cancer subtypes, and only those for which the case was either HCC or GBC were eventually kept. According to the 10th revision of the International Statistical Classification of Diseases, Injury and Causes of Death (ICD10) and the 2nd edition of the International Classification of Diseases for Oncology (ICD-O-2), HCC was defined as C22.0 with morphology codes ICD-O-2 "8170/ 3"and "8180/3", while GBC was defined as tumours in the gallbladder (C23.9), extrahepatic bile ducts (C24.0), ampulla of Vater (C24.1), and biliary tract (C24.8 and C24.9) with morphology code ICD-O-2 "8162/3"1.

Matched case-control pairs of the prostate cancer study were split according to the cancer/tumour stage (for the case). Advanced stage tumours were defined as those with a tumour-node-metastasis (TNM) system score of T3-4 and/or N1-3 and/or M1, or coded as advanced, while localized stage tumours were those with TNM system score of ≤ T2 and N0/x and M0, or stage coded as localized[2].

### 2. Data shared lasso: principle and implementation

We here briefly recall the basic principles of the data shared lasso and provide some details about its implementation in the context of matched case-control studies with multiple subtypes of disease[3–5].

#### a) Principle

Denote by $\beta_{k,j}$ the log-odds ratio representing the mutually adjusted association between metabolite j ($1 \leq j \leq p$) with risk of cancer k ($1 \leq k \leq K$). The data shared lasso[3,4] is based on the following over-parametrization

$$\beta_{k,j} = \mu_j + \delta_{k,j}$$

Here, parameter $\mu_j$ corresponds to the "overall" mutually adjusted association between metabolite $j$ and cancer risk (irrespective of cancer type), while parameter $\delta_{k,j}$ quantifies the type-specific deviation around $\mu_j$ for cancer type $k$, and allows for heterogeneity among the type-specific associations between metabolite $j$ and cancer risk.

Using vector notation, the above parametrization writes $\boldsymbol{\beta}_k = \boldsymbol{\mu} + \boldsymbol{\delta}_k$, with $\boldsymbol{\beta}_k, \boldsymbol{\mu}$ and $\boldsymbol{\delta}_k$ three vectors of size $p$. For an appropriate value of the tuning parameter $\lambda$ (see below), the data shared lasso vector estimate $(\widehat{\boldsymbol{\mu}}, \widehat{\boldsymbol{\delta}}_1, \cdots, \widehat{\boldsymbol{\delta}}_K)$ is a maximizer of the following penalized version of the log-likelihood[5]

$$\sum_{k=1}^{K} L_k(X_k, y_k;\ \boldsymbol{\mu} + \boldsymbol{\delta}_k) - \lambda\left(\|\boldsymbol{\mu}\|_1 + \sum_{k=1}^{K}\|\boldsymbol{\delta}_k\|_1\right)$$

Here, $L_k(X_k, y_k;\ \boldsymbol{\beta}_k)$ denotes the log-likelihood of the conditional logistic regression model computed at the value $\boldsymbol{\beta}_k$ in the matched case-control study corresponding to cancer type $k$ (with design matrix $X_k$and output vector $y_k$), while, for any vector $\boldsymbol{\theta}$ of size $p$, $\|\boldsymbol{\theta}\|_1 = \sum_{j=1}^{p}|\theta_j|$ denotes its $L_1$-norm. For appropriate values of the tuning parameter $\lambda$, and under technical assumptions[4], the data shared lasso allows the identification of metabolites that have a non-null overall association with cancer (those corresponding to non-zero components in $\widehat{\boldsymbol{\mu}}$ ), as well as metabolites whose association with cancer depends on cancer type (those for which the corresponding component of at least one of the vectors $\widehat{\boldsymbol{\delta}}_k$'s is non-null).

**b) Implementation: adaptive version and selection of the tuning parameter $\boldsymbol{\lambda}$**

Data shared lasso estimates were computed using R functions available at https://github.com/NadimBLT/SL1CLR. To enhance sparsity of the vector estimate, we implemented the adaptive version of the data shared lasso[6], under which the estimator is a maximizer of the following penalized criterion

$$\sum_{k=1}^{K} L_k(X_k, y_k;\ \boldsymbol{\mu} + \boldsymbol{\delta}_k) - \lambda\left(\sum_{j=1}^{p}\frac{|\mu_j|}{|\hat{\mu}_j(\lambda_{CV})|_1} + \sum_{k=1}^{K}\sum_{j=1}^{p}\frac{|\delta_{kj}|}{|\hat{\delta}_{kj}(\lambda_{CV})|}\right)$$

where $(\widehat{\boldsymbol{\mu}}(\lambda_{CV}), \widehat{\boldsymbol{\delta}}_1(\lambda_{CV}), \cdots, \widehat{\boldsymbol{\delta}}_K(\lambda_{CV}))$ is an initial data shared lasso estimate computed with tuning parameter selected by cross-validation. The tuning parameter for the adaptive data shared lasso was selected by nested cross-validation[7–9], using the one standard error rule to further enhance sparsity[10].

### 3. Point estimates and confidence intervals

P-values and confidence intervals cannot be directly derived from the data shared lasso. In particular, non-null estimates $\hat{\beta}_{k,j}$ resulting from non-null estimates $\hat{\mu}_j$and $\hat{\delta}_{k,j}$ such that $\hat{\mu}_j\hat{\delta}_{k,j} < 0$ have to be interpreted with caution, especially if $\hat{\beta}_{k,j}$is close to 0. For illustration, consider the example where, for some metabolite $j$, the data shared lasso identifies a positive overall association with cancer ($\hat{\mu}_j = 1$), but also a deviation

from this overall association for some cancer $k$ ($\hat{\delta}_{k,j}$ = -1.05). Then, the estimate of $\beta_{k,j}$ produced by the data shared lasso would be $\hat{\beta}_{k,j}$= -0.05. However, because the $\beta_{k,j}$'s are not penalized, there is no direct way to interpret such results in terms of the nullity of the true parameter $\beta_{k,j}$.
In the present work, we followed the ideas of the ols-hybrid lasso[11] and used standard (*i.e.*, non-penalized) conditional logistic regression models to derive final point estimates and 95% confidence intervals of the non-null $\mu_j$ and $\delta_{k,j}$ identified by the data shared lasso, and eventually the corresponding $\beta_{k,j}$. See paragraph a) below for more details. However, we acknowledge that it corresponds to post-selection inference and the results of this analysis also have to be interpreted with caution as p-values and coverage might not be exact[12].

#### a) Inference under the model identified by the data-shared lasso

Denote by $(\hat{\boldsymbol{\mu}},\ \hat{\boldsymbol{\delta}}_1, \cdots, \hat{\boldsymbol{\delta}}_K)$ the data-shared lasso estimate, and set $J = supp(\hat{\boldsymbol{\mu}})$ and $J_k = supp(\hat{\boldsymbol{\delta}}_k)$ where, for any vector $x$, $supp(x) = \{i \mid x_i \neq 0\}$ denotes its support. Subset $J$ comprises indexes of the metabolites for which an overall association was identified, while $J_k$ corresponds to the indexes of the metabolites for which a deviation from the overall association was identified for the $k$-th cancer type. Then, point estimates and 95% confidence intervals for the non-zero parameters identified by the data shared lasso are derived from conditional logistic regression models based on the following linear predictor

$$\sum_{j \in J} \mu_j \text{met}_j + \sum_{k=1}^{K} \sum_{q \in J_k} \delta_{k,q} \text{met}_q \times \mathbb{1}_k \qquad (1)$$

where met$_j$ denotes the measurement level of metabolite $j$ in the considered sample and $\mathbb{1}_k$ is an indicator variable that equals 1 if the considered sample belongs to the case-control study of the $k$-th cancer type, and 0 otherwise.

For illustration, consider the simple case where the data shared lasso would have identified *(i)* an overall association between cancer and glutamine; *(ii)* an overall association between cancer and proline *(iii)* a deviation from the overall association between cancer and proline for breast cancer; *(iv)* a deviation from the overall null association between cancer and histidine for colorectal cancer. Then we would fit a conditional logistic regression model on the pooled data using linear predictors of the form

$$\mu_{\text{glu}}\text{glutamine} + \left(\mu_{\text{pro}}\text{proline} + \delta_{\text{BrC, pro}}\text{proline} \times \mathbb{1}_{\text{BrC}}\right) + \delta_{\text{CRC, his}}\text{histidine} \times \mathbb{1}_{\text{CRC}} \qquad (2)$$

to derive final point estimates and 95% confidence intervals for $\mu_{\text{glu}}$, $\mu_{\text{pro}}$, $\delta_{\text{BrC, pro}}$, $\delta_{\text{CRC, his}}$ , as well as point estimates and 95% confidence intervals for $\beta_{\text{k, glu}}$, $\beta_{\text{k, pro}}$ and $\beta_{\text{k, his}}$ for all cancer-type *k*.

**b) Inference under "extended" models**

For metabolites in $J$ (*i.e.*, metabolites for which an overall association with cancer was identified by the data shared lasso), the absence of identified cancer type-specific deviations from the overall association could be the result of a lack of statistical power, in particular for HCC and gallbladder and biliary tract cancers where numbers of matched pairs were low. For each metabolite in $J$, we complemented our analysis by considering an "extended" model, derived from model (1) above, but which further allowed fully cancer-type specific associations for that particular metabolite. More specifically, for each $j \in J$, we set $J_{-j} = J \backslash \{j\}$ and $J_{k,-j} = J_k \backslash \{j\}$ for all *k*. Then, for each $j \in J$, we estimated the fully cancer-type specific associations for metabolite $j$ under conditional logistic regression model based on the following linear predictor

$$\sum_{k=1}^{K} \beta_{k,j} \text{met}_j \times \mathbb{I}_k + \sum_{j \in J_{-j}} \mu_j \text{met}_j + \sum_{k=1}^{K} \sum_{q \in J_{k,-j}} \delta_{k,q} \text{met}_q \times \mathbb{I}_k \qquad (3)$$

Cancer type-specific associations for metabolite $j$ are estimated for all cancer types (parameters $\beta_{k,j}$), and the model is adjusted for the associations identified by the data shared lasso for the other metabolites. These point estimates provide information on the possible variability of the association between metabolite $j$ and cancer risk across cancer types. A formal likelihood ratio-test can also be computed to compare models (1) and (3) and assess the statistical significance of the overall observed variability. Again, the p-value of this test has to be interpreted with caution given the post-selection nature of this inference.

**References (Supplementary Material)**

doi:10.1093/biostatistics/kxz063
6. Zou H. The Adaptive Lasso and Its Oracle Properties. *J Am Stat Assoc*. 2006;101(476):1418-1429. doi:10.1198/016214506000000735
7. Krämer N, Schäfer J, Boulesteix AL. Regularized estimation of large-scale gene association networks using graphical Gaussian models. *BMC Bioinformatics*. 2009;10(1):384. doi:10.1186/1471-2105-10-384
8. He K, Wang Y, Zhou X, Xu H, Huang C. An improved variable selection procedure for adaptive Lasso in high-dimensional survival analysis. *Lifetime Data Anal*. 2019;25(3):569-585. doi:10.1007/s10985-018-9455-2
9. Ballout N, Etievant L, Viallon V. On the use of cross-validation for the calibration of the adaptive lasso. *ArXiv200510119 Stat*. Published online July 15, 2021. Accessed December 1, 2021. http://arxiv.org/abs/2005.10119
10. Chen Y, Yang Y. The One Standard Error Rule for Model Selection: Does It Work? *Stats*. 2021;4(4):868-892. doi:10.3390/stats4040051
11. Efron B, Hastie T, Johnstone I, Tibshirani R. Least angle regression. *Ann Stat*. 2004;32(2):407-499. doi:10.1214/009053604000000067
12. Taylor J, Tibshirani R. Post-selection inference for -penalized likelihood models. *Can J Stat*. 2018;46(1):41-61. doi:10.1002/cjs.11313
