## Additional file 2 for "Pan-cancer analysis of pre-diagnostic blood metabolite concentrations in the European Prospective Investigation into Cancer and Nutrition"

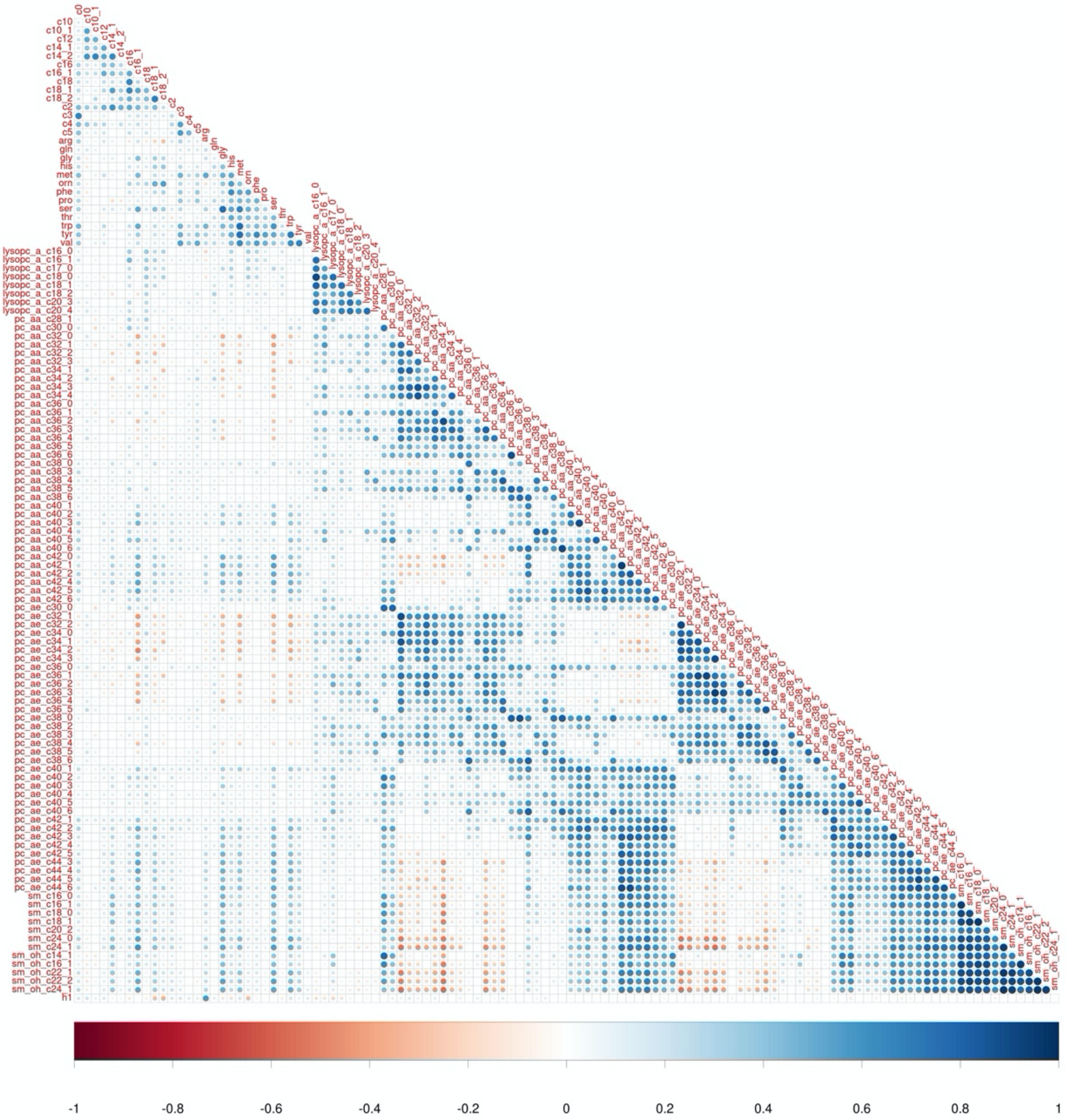


**Figure S1.** Pearson correlation between the 117 metabolites computed in the 5,985 controls of the eight cancer type-specific EPIC studies.

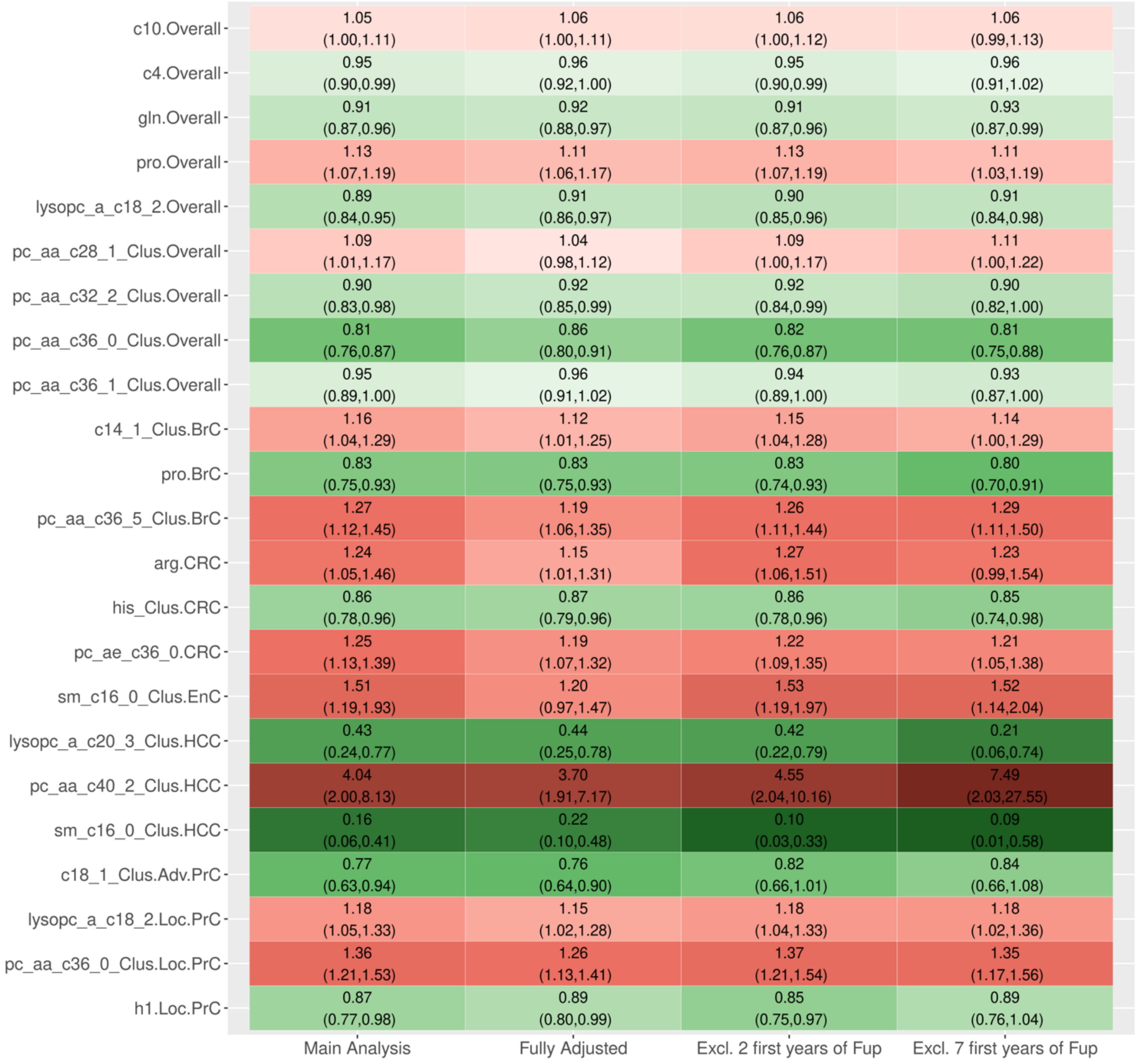


**Figure S2.** Sensitivity analyses of the mutually adjusted odd-ratios for the overall associations and cancer type-specific deviations around the overall association identified by the data shared lasso. Point estimates and confidence intervals were obtained through non-penalized conditional logistic regression models using the design matrix derived from the positions of the non-zero components in the data shared lasso vector estimate $(\hat{\boldsymbol{\mu}},\ \hat{\boldsymbol{\delta}}_1, \cdots, \hat{\boldsymbol{\delta}}_K)$, and based on: *(first column)* residuals of the metabolite measurements after adjustment for BMI (as for the data shared lasso); *(second column)* residuals of the metabolite measurements after adjustment for BMI, education level, waist circumference, height, physical activity, smoking status, alcohol intake, use of non-steroidal anti-inflammatory drugs, and, for women, menopausal status and phase of menstrual cycle; (*third column*) residuals of the metabolite measurements after adjustment for BMI , and excluding pairs for which the case developed cancer within the first two years of follow-up; and (*fourth column*) residuals of the metabolite measurements after adjustment for BMI, and excluding pairs for which the case developed cancer within the first seven years of follow-up. Point estimates and confidence intervals have to be interpreted with caution since they are the result of post-selection inference. In the labels of the y-axis, BrC stands for breast cancer, CRC for colorectal cancer, EnC for endometrial cancer, HCC for hepatocellular carcinoma, and Adv.PrC and Loc.PrC for advanced and localized prostate cancers, respectively. Cluster representatives appear in grey while isolated metabolites appear in black.

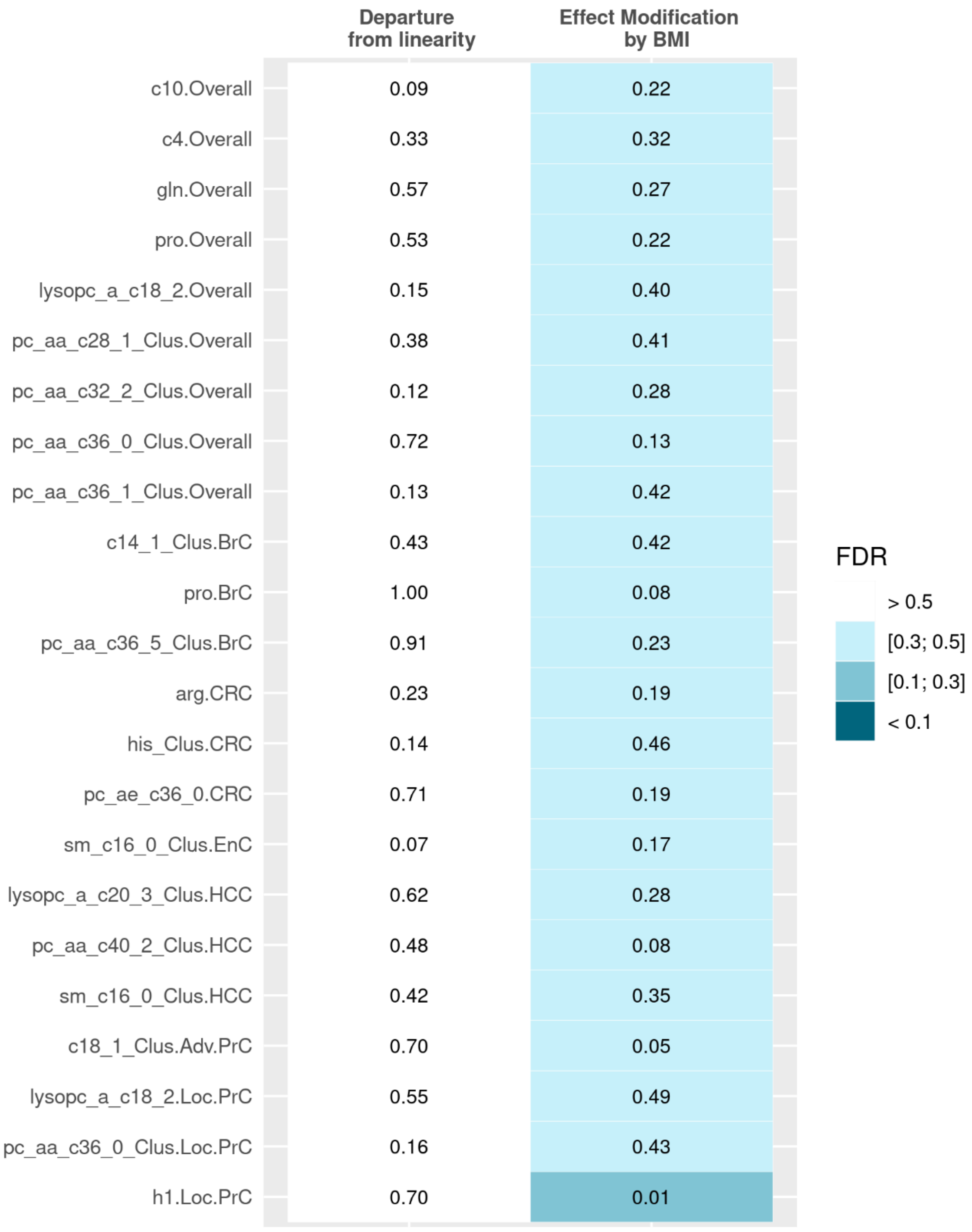


**Figure S3.** P-values of the statistical tests computed to assess *(i)* possible departure from linearity for the associations identified by the data shared lasso; and *(ii)* possible effect modifications by body mass index (BMI) for the associations identified by the data shared lasso. The background colour indicates the value of the corresponding FDR: in particular, none of the FDRs was below 0.10 (the lowest FDR was 0.27). P-values, and FDR, have to be interpreted with caution since they are the result of post-selection inference. In the labels of the y-axis, BrC stands for breast cancer, CRC for colorectal cancer, EnC for endometrial cancer, HCC for hepatocellular carcinoma, and Adv.PrC and Loc.PrC for advanced and localized prostate cancers, respectively.

**ORs ESTIMATED FROM THE MODEL IDENTIFIED BY DATA SHARED LASSO**

| | BrC | CRC | EnC | KiC | GBC | HCC | Adv.PrC | Loc.PrC |
|---|---|---|---|---|---|---|---|---|
| c10 | 1.05 (1.00,1.11) | 1.05 (1.00,1.11) | 1.05 (1.00,1.11) | 1.05 (1.00,1.11) | 1.05 (1.00,1.11) | 1.05 (1.00,1.11) | 1.05 (1.00,1.11) | 1.05 (1.00,1.11) |
| c4 | 0.95 (0.90,0.99) | 0.95 (0.90,0.99) | 0.95 (0.90,0.99) | 0.95 (0.90,0.99) | 0.95 (0.90,0.99) | 0.95 (0.90,0.99) | 0.95 (0.90,0.99) | 0.95 (0.90,0.99) |
| gln | 0.91 (0.87,0.96) | 0.91 (0.87,0.96) | 0.91 (0.87,0.96) | 0.91 (0.87,0.96) | 0.91 (0.87,0.96) | 0.91 (0.87,0.96) | 0.91 (0.87,0.96) | 0.91 (0.87,0.96) |
| pro | 0.94 (0.84,1.05) | 1.13 (1.07,1.19) | 1.13 (1.07,1.19) | 1.13 (1.07,1.19) | 1.13 (1.07,1.19) | 1.13 (1.07,1.19) | 1.13 (1.07,1.19) | 1.13 (1.07,1.19) |
| lysopc_a_c18_2 | 0.89 (0.84,0.95) | 0.89 (0.84,0.95) | 0.89 (0.84,0.95) | 0.89 (0.84,0.95) | 0.89 (0.84,0.95) | 0.89 (0.84,0.95) | 0.89 (0.84,0.95) | 1.06 (0.96,1.17) |
| pc_aa_c28_1_Clus | 1.09 (1.01,1.17) | 1.09 (1.01,1.17) | 1.09 (1.01,1.17) | 1.09 (1.01,1.17) | 1.09 (1.01,1.17) | 1.09 (1.01,1.17) | 1.09 (1.01,1.17) | 1.09 (1.01,1.17) |
| pc_aa_c32_2_Clus | 0.90 (0.83,0.98) | 0.90 (0.83,0.98) | 0.90 (0.83,0.98) | 0.90 (0.83,0.98) | 0.90 (0.83,0.98) | 0.90 (0.83,0.98) | 0.90 (0.83,0.98) | 0.90 (0.83,0.98) |
| pc_aa_c36_0_Clus | 0.81 (0.76,0.87) | 0.81 (0.76,0.87) | 0.81 (0.76,0.87) | 0.81 (0.76,0.87) | 0.81 (0.76,0.87) | 0.81 (0.76,0.87) | 0.81 (0.76,0.87) | 1.11 (1.00,1.23) |
| pc_aa_c36_1_Clus | 0.95 (0.89,1.00) | 0.95 (0.89,1.00) | 0.95 (0.89,1.00) | 0.95 (0.89,1.00) | 0.95 (0.89,1.00) | 0.95 (0.89,1.00) | 0.95 (0.89,1.00) | 0.95 (0.89,1.00) |

**ORs ESTIMATED FROM THE 'EXTENDED' MODELS, ALLOWING FULLY TYPE-SPECIFIC ASSOCIATIONS**

| | BrC | CRC | EnC | KiC | GBC | HCC | Adv.PrC | Loc.PrC |
|---|---|---|---|---|---|---|---|---|
| c10 | 1.10 (0.99,1.24) | 1.05 (0.95,1.17) | 1.03 (0.90,1.17) | 0.92 (0.77,1.10) | 1.11 (0.77,1.59) | 1.34 (0.90,1.99) | 1.12 (0.90,1.39) | 1.02 (0.90,1.16) |
| c4 | 0.96 (0.87,1.06) | 0.93 (0.85,1.01) | 0.90 (0.79,1.01) | 1.02 (0.87,1.20) | 0.88 (0.58,1.34) | 1.10 (0.73,1.66) | 0.90 (0.76,1.06) | 0.98 (0.88,1.08) |
| gln | 0.96 (0.87,1.07) | 0.86 (0.76,0.96) | 0.94 (0.82,1.08) | 0.94 (0.80,1.10) | 0.98 (0.65,1.47) | 0.63 (0.37,1.05) | 0.87 (0.74,1.02) | 0.91 (0.82,1.01) |
| pro | 0.94 (0.85,1.04) | 1.13 (1.03,1.24) | 1.22 (1.07,1.39) | 1.19 (1.02,1.38) | 1.41 (0.89,2.22) | 0.80 (0.46,1.40) | 1.04 (0.90,1.19) | 1.10 (1.00,1.22) |
| lysopc_a_c18_2 | 0.96 (0.85,1.09) | 0.91 (0.82,1.01) | 0.86 (0.74,0.99) | 0.77 (0.65,0.91) | 0.93 (0.62,1.39) | 0.67 (0.35,1.25) | 0.92 (0.78,1.09) | 1.06 (0.95,1.17) |
| pc_aa_c28_1_Clus | 1.04 (0.89,1.21) | 0.97 (0.85,1.11) | 1.20 (0.99,1.47) | 1.08 (0.90,1.31) | 1.13 (0.71,1.81) | 1.67 (0.87,3.23) | 1.16 (0.96,1.38) | 1.16 (1.03,1.32) |
| pc_aa_c32_2_Clus | 0.80 (0.68,0.95) | 0.91 (0.81,1.03) | 0.93 (0.79,1.10) | 0.94 (0.78,1.14) | 0.87 (0.59,1.29) | 0.64 (0.35,1.20) | 0.83 (0.68,1.00) | 0.96 (0.85,1.09) |
| pc_aa_c36_0_Clus | 0.82 (0.71,0.95) | 0.82 (0.73,0.91) | 0.77 (0.66,0.90) | 0.85 (0.72,0.99) | 0.54 (0.34,0.86) | 0.93 (0.58,1.49) | 0.86 (0.74,1.00) | 1.11 (1.00,1.23) |
| pc_aa_c36_1_Clus | 0.85 (0.75,0.98) | 0.99 (0.89,1.11) | 0.88 (0.76,1.02) | 1.10 (0.94,1.28) | 0.92 (0.63,1.34) | 1.52 (0.83,2.81) | 0.87 (0.75,1.01) | 0.96 (0.87,1.05) |

**Figure S4.** Association (odds-ratio) with risks of the eight cancer types for the nine metabolites that had an overall association with cancer risk in our main analysis. (*Top*) Point estimates and 95% confidence intervals were obtained through non-penalized conditional logistic regression models using the design matrix derived from the positions of the non-zero components in the data shared lasso vector estimate $(\hat{\boldsymbol{\mu}},\ \hat{\boldsymbol{\delta}}_1, \cdots, \hat{\boldsymbol{\delta}}_K)$; see Section 3.a) in the Supplementary Material for details. (*Bottom*) For each of the nine features, point estimates and 95% confidence intervals were obtained through non-penalized conditional logistic regression models using the design matrix derived from the positions of the non-zero components in the data shared lasso vector estimate $(\hat{\boldsymbol{\mu}},\ \hat{\boldsymbol{\delta}}_1, \cdots, \hat{\boldsymbol{\delta}}_K)$, but further allowing for fully type-specific associations between that particular feature and cancer risk; see Section 3.b) in in the Supplementary Material for details. The p-values of the likelihood ratio tests comparing the two types of models were 0.65, 0.85, 0.63, 0.42, 0.39, 0.32, 0.53, 0.55 and 0.11 for c10, c4, gln, pro, lysopc_a_c18_2, pc_aa_c28_1_Clus, pc_aa_c32_2_Clus, pc_aa_c36_0_Clus and pc_aa_c36_1_Clus, respectively, suggesting the absence of type-specific deviations beyond those identified by the data shared lasso for these nine metabolites. Point estimates, confidence intervals and p-values have to be interpreted with caution since they are the result of post-selection inference. In the column labels, BrC stands for breast cancer, CRC for colorectal cancer, EnC for endometrial cancer, KiC for Kidney cancer, GBC for gallbladder and biliary tract cancer, HCC for hepatocellular carcinoma, and Adv.PrC and Loc.PrC for advanced and localized prostate cancers, respectively. Cluster representatives appear in grey.

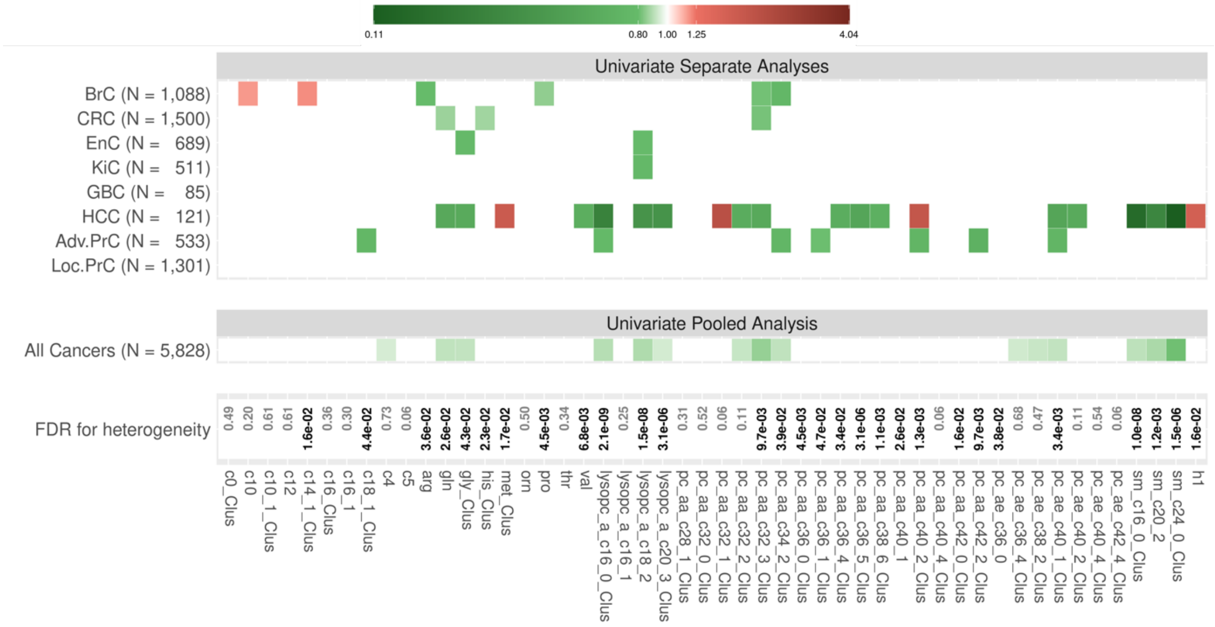


**Figure S5.** Summary of the results from our univariate analyses: *(top)* associations with an FDR < 5% in the univariate analyses of the association between each feature and risk of each cancer type in conditional logistic regression models adjusted for BMI; *(center)* associations with an FDR < 5% in the univariate analyses of the association between each feature and the risk of cancer (after pooling the data from the eight cancer-type specific studies together, thus ignoring cancer types), in conditional logistic regression models adjusted for BMI; *(bottom)* FDR for the statistical test of a heterogeneity among its (non-mutually adjusted) type-specific associations. In the top and center panels, white entries correspond to the absence of identified associations, while green and red entries correspond to inverse and positive associations, respectively. The more intense the colour, the larger the absolute value of the log-odds-ratio). The x-axis represents the 50 features (33 cluster representatives and 17 isolated metabolites). In the labels of the y-axis, numbers correspond to numbers of pairs for each type-specific cancer (and in total for the pooled analysis), while BrC stands for breast cancer, CRC for colorectal cancer, EnC for endometrial cancer, KiC for Kidney cancer, GBC for gallbladder and biliary tract cancer, HCC for hepatocellular carcinoma, and Adv.PrC and Loc.PrC for advanced and localized prostate cancers, respectively.

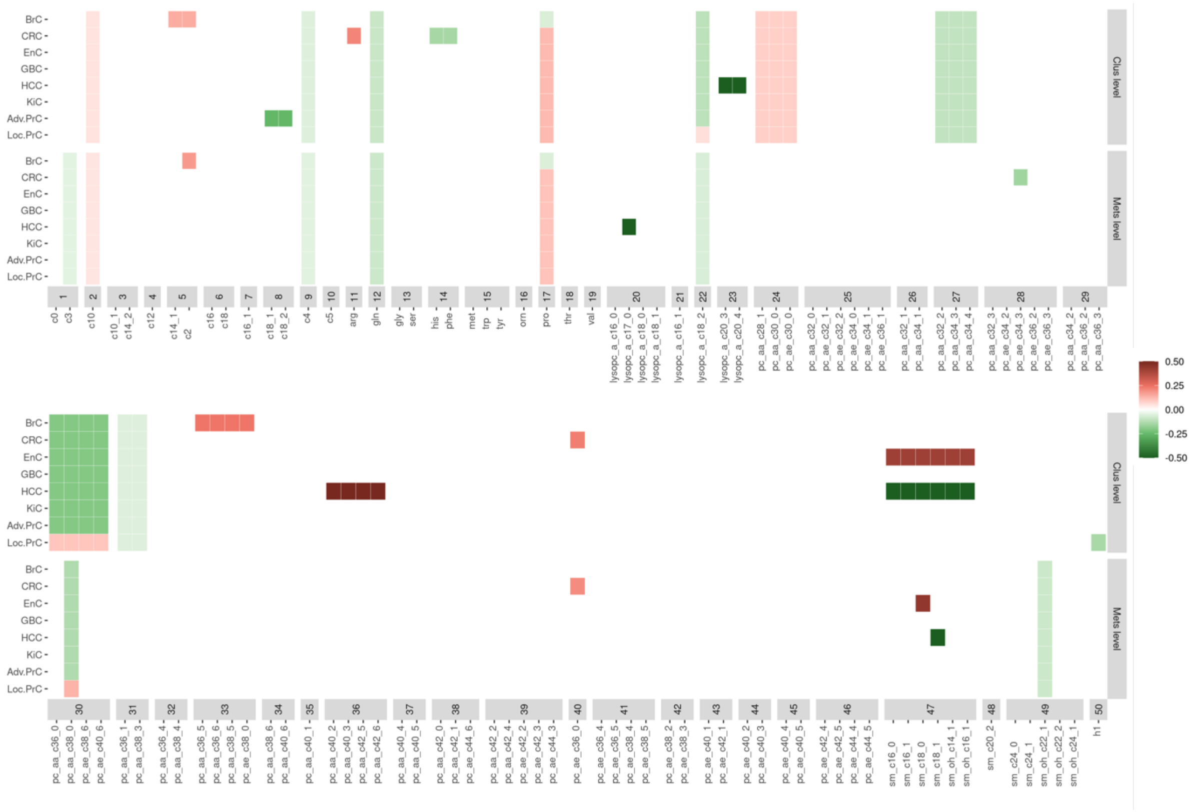


**Figure S6.** Comparison of the associations identified by the data shared lasso when working with the 50 features (Clus Level, as in our main analysis) or with the original 117 metabolites (Mets Level). The 117 metabolites are organized by cluster, and for the associations identified when working at the cluster level, the figure displays this association for all the metabolites that compose that cluster. Overall, the analysis conducted at the cluster level identified more associations, but most results from the two types of analyses were consistent. For lysoPCs for example, the two analyses did not identify exactly the same associations, but they both suggested *(i)* an overall inverse association with cancer risk and *(ii)* a stronger inverse association with HCC (the analysis conducted at the cluster level also suggested a weaker association with localized PrC). In the labels of the *y* axis, BrC stands for breast cancer, CRC for colorectal cancer, EnC for endometrial cancer, KiC for Kidney cancer, GBC for gallbladder and biliary tract cancer, HCC for hepatocellular carcinoma, and Adv.PrC and Loc.PrC for advanced and localized prostate cancers, respectively.

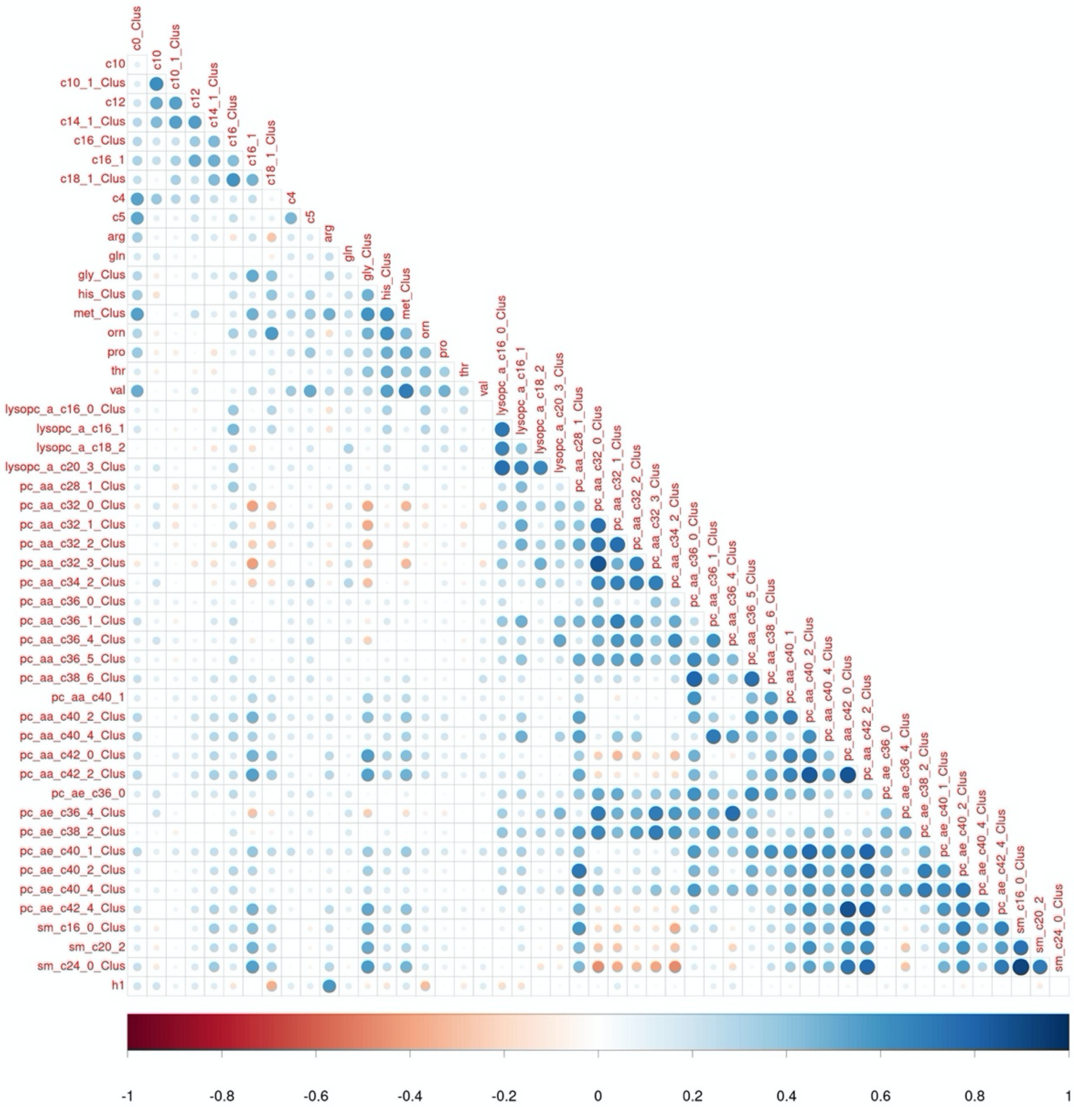


**Figure S7.** Pearson correlation between the 50 features computed in the 5,828 controls of the eight cancer type-specific EPIC studies. The 50 features correspond to 17 isolated metabolites and 33 representatives of clusters of strongly-correlated metabolites, obtained by hierarchical clustering of the 117 metabolites among the 5,828 controls. Clusters are labelled as "metabo_Clus", with "metabo" being one of the metabolites that compose that cluster.

**Table S1.** List of the 117 metabolites studied in the main analysis, and the 16 additional metabolites studied in the sensitivity analysis, following the exclusion of the participants from the second CRC study.

| Class | SubClass | Compounds | Name |
|---|---|---|---|
| **Metabolites used in the main analysis** | | | |
| acylcarnitines | acylcarnitines | C0 | Carnitine |
| acylcarnitines | acylcarnitines | C10 | Decanoylcarnitine |
| acylcarnitines | acylcarnitines | C10_1 | Decenoylcarnitine |
| acylcarnitines | acylcarnitines | C12 | Dodecanoylcarnitine |
| acylcarnitines | acylcarnitines | C14_1 | Tetradecenoylcarnitine |
| acylcarnitines | acylcarnitines | C14_2 | Tetradecadienylcarnitine |
| acylcarnitines | acylcarnitines | C16 | Hexadecanoylcarnitine |
| acylcarnitines | acylcarnitines | C16_1 | Hexadecenoylcarnitine |
| acylcarnitines | acylcarnitines | C18 | Octadecanoylcarnitine |
| acylcarnitines | acylcarnitines | C18_1 | Octadecenoylcarnitine |
| acylcarnitines | acylcarnitines | C18_2 | Octadecadienylcarnitine |
| acylcarnitines | acylcarnitines | C2 | Acetylcarnitine |
| acylcarnitines | acylcarnitines | C3 | Propionylcarnitine |
| acylcarnitines | acylcarnitines | C4 | Butyrylcarnitine |
| acylcarnitines | acylcarnitines | C5 | Valerylcarnitine |
| aminoacids | non-essential aa | Arg | Arginine |
| aminoacids | non-essential aa | Gln | Glutamine |
| aminoacids | non-essential aa | Gly | Glycine |
| aminoacids | essential aa | His | Histidine |
| aminoacids | essential aa | Met | Methionine |
| aminoacids | non-essential aa | Orn | Ornithine |
| aminoacids | essential aa | Phe | Phenylalanine |
| aminoacids | non-essential aa | Pro | Proline |
| aminoacids | non-essential aa | Ser | Serine |
| aminoacids | essential aa | Thr | Threonine |
| aminoacids | essential aa | Trp | Tryptophan |
| aminoacids | non-essential aa | Tyr | Tyrosine |
| aminoacids | essential aa | Val | Valine |
| glycerophospholipids | saturated | lysoPC_a_C16_0 | lysoPC a C16:0 |
| glycerophospholipids | mono-unsaturated | lysoPC_a_C16_1 | lysoPC a C16:1 |
| glycerophospholipids | saturated | lysoPC_a_C17_0 | lysoPC a C17:0 |
| glycerophospholipids | saturated | lysoPC_a_C18_0 | lysoPC a C18:0 |
| glycerophospholipids | mono-unsaturated | lysoPC_a_C18_1 | lysoPC a C18:1 |
| glycerophospholipids | poly-unsaturated | lysoPC_a_C18_2 | lysoPC a C18:2 |
| glycerophospholipids | poly-unsaturated | lysoPC_a_C20_3 | lysoPC a C20:3 |

| | | | |
|---|---|---|---|
| glycerophospholipids | poly-unsaturated | lysoPC_a_C20_4 | lysoPC a C20:4 |
| glycerophospholipids | mono-unsaturated | PC_aa_C28_1 | PC aa C28:1 |
| glycerophospholipids | saturated | PC_aa_C30_0 | PC aa C30:0 |
| glycerophospholipids | saturated | PC_aa_C32_0 | PC aa C32:0 |
| glycerophospholipids | mono-unsaturated | PC_aa_C32_1 | PC aa C32:1 |
| glycerophospholipids | poly-unsaturated | PC_aa_C32_2 | PC aa C32:2 |
| glycerophospholipids | poly-unsaturated | PC_aa_C32_3 | PC aa C32:3 |
| glycerophospholipids | mono-unsaturated | PC_aa_C34_1 | PC aa C34:1 |
| glycerophospholipids | poly-unsaturated | PC_aa_C34_2 | PC aa C34:2 |
| glycerophospholipids | poly-unsaturated | PC_aa_C34_3 | PC aa C34:3 |
| glycerophospholipids | poly-unsaturated | PC_aa_C34_4 | PC aa C34:4 |
| glycerophospholipids | saturated | PC_aa_C36_0 | PC aa C36:0 |
| glycerophospholipids | mono-unsaturated | PC_aa_C36_1 | PC aa C36:1 |
| glycerophospholipids | poly-unsaturated | PC_aa_C36_2 | PC aa C36:2 |
| glycerophospholipids | poly-unsaturated | PC_aa_C36_3 | PC aa C36:3 |
| glycerophospholipids | poly-unsaturated | PC_aa_C36_4 | PC aa C36:4 |
| glycerophospholipids | poly-unsaturated | PC_aa_C36_5 | PC aa C36:5 |
| glycerophospholipids | poly-unsaturated | PC_aa_C36_6 | PC aa C36:6 |
| glycerophospholipids | saturated | PC_aa_C38_0 | PC aa C38:0 |
| glycerophospholipids | poly-unsaturated | PC_aa_C38_3 | PC aa C38:3 |
| glycerophospholipids | poly-unsaturated | PC_aa_C38_4 | PC aa C38:4 |
| glycerophospholipids | poly-unsaturated | PC_aa_C38_5 | PC aa C38:5 |
| glycerophospholipids | poly-unsaturated | PC_aa_C38_6 | PC aa C38:6 |
| glycerophospholipids | mono-unsaturated | PC_aa_C40_1 | PC aa C40:1 |
| glycerophospholipids | poly-unsaturated | PC_aa_C40_2 | PC aa C40:2 |
| glycerophospholipids | poly-unsaturated | PC_aa_C40_3 | PC aa C40:3 |
| glycerophospholipids | poly-unsaturated | PC_aa_C40_4 | PC aa C40:4 |
| glycerophospholipids | poly-unsaturated | PC_aa_C40_5 | PC aa C40:5 |
| glycerophospholipids | poly-unsaturated | PC_aa_C40_6 | PC aa C40:6 |
| glycerophospholipids | saturated | PC_aa_C42_0 | PC aa C42:0 |
| glycerophospholipids | mono-unsaturated | PC_aa_C42_1 | PC aa C42:1 |
| glycerophospholipids | poly-unsaturated | PC_aa_C42_2 | PC aa C42:2 |
| glycerophospholipids | poly-unsaturated | PC_aa_C42_4 | PC aa C42:4 |
| glycerophospholipids | poly-unsaturated | PC_aa_C42_5 | PC aa C42:5 |
| glycerophospholipids | poly-unsaturated | PC_aa_C42_6 | PC aa C42:6 |
| glycerophospholipids | saturated | PC_ae_C30_0 | PC ae C30:0 |
| glycerophospholipids | mono-unsaturated | PC_ae_C32_1 | PC ae C32:1 |
| glycerophospholipids | poly-unsaturated | PC_ae_C32_2 | PC ae C32:2 |

| | | | |
|---|---|---|---|
| glycerophospholipids | saturated | PC_ae_C34_0 | PC ae C34:0 |
| glycerophospholipids | mono-unsaturated | PC_ae_C34_1 | PC ae C34:1 |
| glycerophospholipids | poly-unsaturated | PC_ae_C34_2 | PC ae C34:2 |
| glycerophospholipids | poly-unsaturated | PC_ae_C34_3 | PC ae C34:3 |
| glycerophospholipids | saturated | PC_ae_C36_0 | PC ae C36:0 |
| glycerophospholipids | mono-unsaturated | PC_ae_C36_1 | PC ae C36:1 |
| glycerophospholipids | poly-unsaturated | PC_ae_C36_2 | PC ae C36:2 |
| glycerophospholipids | poly-unsaturated | PC_ae_C36_3 | PC ae C36:3 |
| glycerophospholipids | poly-unsaturated | PC_ae_C36_4 | PC ae C36:4 |
| glycerophospholipids | poly-unsaturated | PC_ae_C36_5 | PC ae C36:5 |
| glycerophospholipids | saturated | PC_ae_C38_0 | PC ae C38:0 |
| glycerophospholipids | poly-unsaturated | PC_ae_C38_2 | PC ae C38:2 |
| glycerophospholipids | poly-unsaturated | PC_ae_C38_3 | PC ae C38:3 |
| glycerophospholipids | poly-unsaturated | PC_ae_C38_4 | PC ae C38:4 |
| glycerophospholipids | poly-unsaturated | PC_ae_C38_5 | PC ae C38:5 |
| glycerophospholipids | poly-unsaturated | PC_ae_C38_6 | PC ae C38:6 |
| glycerophospholipids | mono-unsaturated | PC_ae_C40_1 | PC ae C40:1 |
| glycerophospholipids | poly-unsaturated | PC_ae_C40_2 | PC ae C40:2 |
| glycerophospholipids | poly-unsaturated | PC_ae_C40_3 | PC ae C40:3 |
| glycerophospholipids | poly-unsaturated | PC_ae_C40_4 | PC ae C40:4 |
| glycerophospholipids | poly-unsaturated | PC_ae_C40_5 | PC ae C40:5 |
| glycerophospholipids | poly-unsaturated | PC_ae_C40_6 | PC ae C40:6 |
| glycerophospholipids | mono-unsaturated | PC_ae_C42_1 | PC ae C42:1 |
| glycerophospholipids | poly-unsaturated | PC_ae_C42_2 | PC ae C42:2 |
| glycerophospholipids | poly-unsaturated | PC_ae_C42_3 | PC ae C42:3 |
| glycerophospholipids | poly-unsaturated | PC_ae_C42_4 | PC ae C42:4 |
| glycerophospholipids | poly-unsaturated | PC_ae_C42_5 | PC ae C42:5 |
| glycerophospholipids | poly-unsaturated | PC_ae_C44_3 | PC ae C44:3 |
| glycerophospholipids | poly-unsaturated | PC_ae_C44_4 | PC ae C44:4 |
| glycerophospholipids | poly-unsaturated | PC_ae_C44_5 | PC ae C44:5 |
| glycerophospholipids | poly-unsaturated | PC_ae_C44_6 | PC ae C44:6 |
| sphingolipids | sphingomyelin | SM_OH_C14_1 | SM (OH) C14:1 |
| sphingolipids | sphingomyelin | SM_OH_C16_1 | SM (OH) C16:1 |
| sphingolipids | sphingomyelin | SM_OH_C22_1 | SM (OH) C22:1 |
| sphingolipids | sphingomyelin | SM_OH_C22_2 | SM (OH) C22:2 |
| sphingolipids | sphingomyelin | SM_OH_C24_1 | SM (OH) C24:1 |
| sphingolipids | sphingomyelin | SM_C16_0 | SM C16:0 |
| sphingolipids | sphingomyelin | SM_C16_1 | SM C16:1 |

| | | | |
|---|---|---|---|
| sphingolipids | sphingomyelin | SM_C18_0 | SM C18:0 |
| sphingolipids | sphingomyelin | SM_C18_1 | SM C18:1 |
| sphingolipids | sphingomyelin | SM_C20_2 | SM C20:2 |
| sphingolipids | sphingomyelin | SM_C24_0 | SM C24:0 |
| sphingolipids | sphingomyelin | SM_C24_1 | SM C24:1 |
| sugars | sugars | H1 | Sum of Hexoses |
| **Additional metabolites used in the analysis following the exclusion of the second CRC sub-study** | | | |
| aminoacids | non-essential aa | Ala | Alanine |
| aminoacids | non-essential aa | Asn | Asparagine |
| aminoacids | non-essential aa | Asp | Aspartate |
| aminoacids | non-essential aa | Cit | Citrulline |
| aminoacids | non-essential aa | Glu | Glutamate |
| aminoacids | essential aa | Ile | Isoleucine |
| aminoacids | essential aa | Leu | Leucine |
| aminoacids | essential aa | Lys | Lysine |
| biogenic amines | biogenic amines | alpha_AAA | alpha-Aminoadipic acid |
| biogenic amines | biogenic amines | Creatinine | Creatinine |
| biogenic amines | biogenic amines | Kynurenine | Kynurenine |
| biogenic amines | biogenic amines | Serotonin | Serotonin |
| biogenic amines | biogenic amines | Spermidine | Spermidine |
| biogenic amines | biogenic amines | Spermine | Spermine |
| biogenic amines | biogenic amines | Taurine | Taurine |
| glycerophospholipids | poly-unsaturated | PC_ae_C30_2 | PC ae C30:2 |
